# Maternal caffeine consumption during pregnancy and offspring cord blood DNA methylation: a meta-analysis of epigenome-wide association studies

**DOI:** 10.1101/2023.04.26.23288858

**Authors:** Laura Schellhas, Giulietta S Monasso, Janine F Felix, Vincent WV Jaddoe, Peiyuan Huang, Sílvia Fernández-Barrés, Martine Vrijheid, Giancarlo Pesce, Isabella Annesi-Maesano, Christian M Page, Anne-Lise Brantsæter, Mona Bekkhus, Siri E Håberg, Stephanie J London, Marcus R Munafò, Luisa Zuccolo, Gemma C Sharp

## Abstract

**Background:** Women are advised to limit caffeine consumption during pregnancy. To uncover the potential epigenetic effects of intrauterine caffeine exposure, we investigated the association of maternal caffeine consumption during pregnancy with genome-wide DNA methylation in cord blood.

**Methods:** We meta-analysed results from epigenome-wide methylation studies at individual probes and in differentially methylated region (DMR) analysis across 6 European pregnancy and birth cohorts (ALSPAC, BiB, MoBa, Generation R, INMA, EDEN; total n = 3742). Methylation was assessed with lllumina Infinium 450k or EPIC arrays. Maternal caffeine consumption (mg/day) from coffee, tea and cola was derived from questionnaires between weeks 12 - 22 of pregnancy. We investigated associations of methylation with overall and beverage-specific caffeine intake in models adjusted for maternal education, age, BMI, smoking during pregnancy, parity, cord-blood cell proportions and 20 surrogate variables.

**Results:** One CpG site (cg19370043, nearest gene *PRRX1*) was associated with maternal caffeine consumption after FDR adjustment for multiple testing and one CpG sites (cg14591243, nearest gene *STAG1*) was associated with maternal cola consumption. We found evidence for 12-22 DMRs for each of the caffeine models but little overlap between DMRs identified for specific caffeinated beverages.

**Conclusions:** In models adjusted for maternal smoking and other potential confounders, we found little evidence to support an intrauterine effect of caffeine on offspring DNA methylation. Comparing associations across different sources of caffeine provided no evidence for caffeine being the causal agent. It is possible that our study did not have adequate statistical power to detect very small associations between maternal caffeine and offspring DNA methylation.

**Key messages:**

- This large-scale meta-analysis of epigenomewide association studies across six European cohorts does not support an intrauterine effect of caffeine on offspring cord blood DNA methylation.
- Lack of overlap between associations with different caffeinated drinks suggest that any (weak) associations were driven by diverse confounding structures of different caffeinated drinks, rather than caffeine per se.
- More research is needed to understand the biological mechanisms driving potential effects of caffeine on offspring health.

## Introduction

The effects of intrauterine caffeine exposure on offspring health outcomes are not well understood. And as a result, pregnancy guidelines for caffeine use vary during pregnancy (1). Throughout pregnancy, the metabolic rate of caffeine gradually decreases. In the second and third trimester of pregnancy the half-life of caffeine can be up until 18 hours, which is about four times longer than outside of pregnancy (2). Due to pregnancy-related changes to the caffeine metabolism and the potential of caffeine readily crossing the placenta barrier, the European Food Safety Authority (EFSA) states *“…unborn children to be the most vulnerable group for adverse effects of caffeine among the general population”* (*3*). Current pregnancy guidelines for caffeine consumption across Asia, Oceania, Europe, Latin America, and North America recommend avoiding or limiting caffeine intake during pregnancy (1). These guidelines are based evidence from observational study designs, with results likely affected by selection, measurement, and confounding biases and therefore cannot differentiate confounded from causal effects (4). This is especially problematic as caffeine consumption is strongly culturally bound, and the sources of caffeine may vary between different countries and be differentially socially patterned (1,5–7).

Identification of a molecular pathway that could explain how prenatal exposure to caffeine potentially increases the risk for adverse offspring health outcomes would add to the evidence base and strengthen the causal interpretation of these associations. DNA methylation in particular has been proposed as the mediating mechanism between prenatal exposures and offspring health outcomes (8). In this epigenetic modification, a methyl group is added to a cytosine-phosphate-guanine (CpG) site in the genome (9). A recent epigenome-wide association study (EWAS) meta-analysis of adult populations found coffee consumption to be associated with own peripheral blood DNA methylation at 11 CpG sites (10). No published studies have examined the role of prenatal exposure to caffeine in humans, but there is evidence from animal studies that prenatal caffeine consumption can lead to DNA methylation changes and offspring risk of disease (11–14).

In this study, we explored the association between maternal caffeine consumption (from tea, coffee, and cola) during pregnancy and offspring cord-blood DNA methylation across the genome, by meta-analysing results from six international pregnancy and birth cohorts from the Pregnancy and Childhood Epigenetics (PACE) consortium (8).

## Methods

### Meta-analysis of epigenome-wide association studies (EWAS)

#### Participating cohorts

The EWAS meta-analysis included six independent prospective pregnancy and birth cohorts from the PACE consortium (8), that had data available on cord blood DNA methylation and maternal caffeine consumption during pregnancy. The total sample (N = 3,731) included two UK based cohorts (ALSPAC (15,16) and BiB)(17,18), one Dutch (Generation R) (19,20), one Norwegian (MoBa1) (21), one Spanish (INMA) (22), and one French (EDEN) (23)cohort. Recruitment periods varied by cohort and took place between the beginning of the 1990s (ALSPAC) and 2008 (MoBa1). Ethical approval was obtained by local ethics committees and informed consent for the use of data was obtained from all participants. More details about the individual cohorts can be found in the supplementary information.

#### Measurement of maternal caffeine intake during pregnancy

Assessment of maternal caffeine consumption varied by cohort and is described in more detail in Supplementary Information. Generally, mothers self-reported the number of cups they consumed of caffeinated coffee, tea, and cola in questionnaires between week 12 to 22 of pregnancy. All cohorts used Food Frequency Questionnaires (FFQ) (24) except for Generation R, which also did not have information on caffeinated cola consumption available. Cups per day were transformed to milligrams of caffeine per day (mg/day), based on the assumption that one standard sized cup of coffee contains 57 mg, one cup of tea contains 27 mg, and one cup of cola contains 20 mg of caffeine (25). A continuous total caffeine score was calculated by summing the caffeine content from each caffeinated drink in mg/day (supplementary information). In addition to the continuous score, we investigated whether any caffeine exposure (regardless of the amount of caffeine) during pregnancy might have an effect on offspring DNA methylation, and thus dichotomised total caffeine into 0 mg/day = none, and > 0 mg/day = any.

#### Measurement of DNA methylation

Cohorts assessed cord blood DNA methylation data individually, using their own laboratory methods, quality control, and normalisation. DNA methylation data was sampled using the lllumina Infinium® HumanMethylation450 (486,425 probes), except for BiB, which used the Illumina EPIC BeadChip array (866,553 probes). Probes on single nucleotide polymorphisms (SNPs), cross-hybridizing probes (26), and probes on the sex chromosomes were excluded. In the final meta-analysis, only probes that were available in both arrays (maximum 364,678) were included. Methylation was measured using normalised beta values ranging from 0 to 1, representing 0 to 100% methylation.

#### Covariates

To adjust for variation in DNA methylation driven by cell composition, models were adjusted for cell proportions estimated using the Houseman method with a cord blood reference panel (27,28). Offspring sex was used to conduct sex-stratified sensitivity analyses because sex-specific DNA methylation differences can still be observed even when restricting analyses to autosomes and removing probes that are cross-reactive with sex chromosomes (29). Another sensitivity analysis was conducted where the unstratified models were additionally adjusted for gestational age at birth. Gestational age could be a mediating factor as it is robustly associated with DNA methylation (30,31) and there is some evidence that it can be associated with prenatal caffeine exposures (32,33). To avoid introducing collider bias, we adjusted for gestational age in separate instead of the main models (34). To adjust for possible technical variation, all cohorts generated 20 surrogate variables and included them in models - as is standard practice in the field (35).

Each model contained the following covariates (35,36): An ordinal measure representing maternal education as a proxy for socio-economic position, maternal age in years, maternal BMI (kg/m^2^), a binary measure of maternal smoking during pregnancy (e.g., in ALSPAC: 0 = no smoking or giving up smoking during the first trimester, 1 = smoking after the 1st trimester), and a binary assessment of parity (1= one or more previous children, 0 = no previous children). See supplementary information for the classifications of covariates in each cohort.

### Statistical analyses

#### Cohort-specific statistical analyses

##### Probe-level analysis

The analysis plan and R-script is available on GitHub (https://github.com/ammegandchips/Prenatal_Caffeine). Cohorts were asked to exclude multiple pregnancies (e.g., twins) and siblings so that each mother was only included once in the dataset. If cohorts included more than one major ethnic group, they were asked to run the EWAS analysis separately for each group. The EWAS R script included: a function to remove probes classified as an outlier according to the Tukey method of outlier removal (values⍰<⍰25th percentile⍰−⍰3*interquartile range and values⍰>⍰75th percentile⍰+⍰3*interquartile range)(37), a function to generate surrogate variables using the R package SVA (38), and a function to run an EWAS of each model using the R package Limma (39). In a second sensitivity analysis, the binary (any vs. none) and total continuous unstratified models were additionally adjusted for gestational age at birth. In a separate sensitivity analysis without gestational age adjustment, the binary (any vs. none) and total continuous caffeine models were stratified by offspring’s sex. For quality assurance, an independent shadow meta-analysis was conducted by a co-author of the University of Bristol (Peiyuan Huang).

Prior to meta-analysing summary results from each cohort, quality checks were conducted to ensure that the EWAS were properly conducted and there were no problems with the data, in line with standard practice in the field (35,40) (supplementary information).

#### Differentially methylated regions (DMR)

The probe level approach was complemented using a regional analysis, which considers DNA methylation at clusters of neighbouring CpG sites throughout the epigenome. This approach is more statistically powerful and arguably more biologically plausible; neighbouring CpG sites are assumed to exert similar biological functions. We used the DMRff method (41) to identify differentially methylated regions (DMRs). Cohorts were supplied with an R script to conduct the DMR analysis using their own data (https://github.com/ammegandchips/Prenatal_Caffeine/blob/master/dmrff.mat.caff.EWAS.cohorts.r). Probes were annotated to the human reference genome version 19, build 37h using the annotation data available from the R-package *meffil* (42).

#### Meta-analysis

##### Probe level meta-analysis

Results were meta-analysed with fixed effect estimates weighted by the inverse of the variance using the software METAL (43). Multiple testing was accounted for using a 5% false discovery rate (FDR) (44). Meta-analysed results were scrutinised in a similar manner as the individual cohort results and leave-one-cohort-out analysis using the R package *metafor* (*45*) was performed on the CpGs that showed evidence to be associated with maternal caffeine consumption. Results were deemed to be driven by a single cohort (and therefore to “fail” the leave-one-out test), if the meta-analysis effect estimate changed direction, moved towards the null by more than 20%, or had a confidence interval that included 0 after removal of a single cohort.

##### DMR meta-analysis

DMR cohort results were meta-analysed using an inverse-variance weighted fixed effects approach using the *dmrff.meta* function in the DMRFF R package (41). We defined a DMR as a region with at least two CpG sites with the same direction of effect and a Bonferroni adjusted P-value (P_Bonferroni_) < 0.05.

##### Causal inference and sensitivity analyses

Beverage-specific effects of the meta-analysed probe-level and DMR results were investigated by comparing the congruence between associations found using different sources of caffeine (that could have different confounding structures). Whereas high congruence between results of the different caffeine models (in terms of CpG site hits and/or genes annotated to CpG sites found in each model) would provide evidence for caffeine being the causal agent driving effects, beverage-specific effects would indicate that factors other than caffeine are driving associations.

To find out which gene pathways are linked to the CpG sites of the caffeine-associated DMRs, a gene ontology analysis was run using the R package *missMethyl* (46). We tested enrichment of gene ontology (GO) categories and Kyoto Encyclopaedia of Genes and Genomes (KEGG) pathways.

## Results

### Sample characteristics

#### Maternal caffeine consumption during pregnancy

In all cohorts, most mothers (80-94%) consumed at least some caffeine during weeks 1 to 28 of gestation, with a weighted mean of 85 mg/day over all cohorts, but with large variation within and between cohorts (weighted average SD = 82 mg/day, Table 1). Approximately 14% of mothers in the total sample consumed more caffeine than the commonly recommended caffeine limit of 200 mg/day. Across all cohorts, coffee and tea were the most common sources of caffeine, with coffee being the most common source in all cohorts except for the United Kingdom based cohorts ALSPAC and BiB, where the most common source was caffeinated tea (Table 1).

**Table 1.** Overview of daily maternal caffeine consumption during pregnancy in the individual cohorts.

| Cohort (N) | Weeks of gestation of dietary assessment | Mean total daily caffeine intake (SD) | N users (%) > 200 mg of daily caffeine intake* | Mean daily intake of coffee (SD) | Mean daily intake of tea (SD) | Mean daily intake of cola (SD) |
| --- | --- | --- | --- | --- | --- | --- |
| **ALSPAC**<br>(n = 729) | 18 | 135 (94) | 197 (27%) | 52 (69) | 72 (58) | 3 (6) |
| **BIB**<br>(Asian; n = 353) | 26-28 | 49 (47) | 5 (2%) | 11 (33) | 41 (34) | 12 (20) |
| **BIB**<br>(white EU; n = 306) | 26-28 | 112 (105) | 50 (19%) | 46 (65) | 66 (63) | 15 (22) |
| **Generation R**<br>(n = 798) | 18-25 | 115 (96) | 132 (20%) | 118 (78) | 57 (62) | Not available |
| **INMA**<br>(n = 378) | 12 | 111 (129) | 26 (8%) | 79 (120) | 25 (46) | 7 (12) |
| **EDEN**<br>(n = 156) | 24-28 | 37 (43) | 1 (<1%) | 25 (42) | 6 (15) | 6 (9) |
| **MoBa1**<br>(N = 999) | 22 | 105(106) | 108 (11%) | 70 (106) | 18 (27) | 14 (27) |
| **Total and % or M and SD\*\***<br>(n = 3,724) | - | 85 (82) | 519 (14%) | 46 (65) | 26 (35) | 5 (11) |

*Note. M = Mean. SD = Standard deviation. \* Mothers were grouped as users of caffeine if they indicated to consume more than zero cups of coffee, tea or cola. caffeine content in milligrams per day. \*\* In the Total row, average caffeine content was calculated by weighting by the inverse variance for each cohort. EU = European.*

#### Demographics

A general overview over the demographics of the individual cohorts can be found in Table 2. Except for mothers from BiB, cohorts included slightly more mothers with higher (high school diploma or above) than lower educational attainment. Around 15% of mothers smoked after the 2^nd^ trimester of pregnancy. Mothers who consumed caffeine during pregnancy were about twice as likely to have smoked during pregnancy (20%), compared to mothers who did not consume caffeine during pregnancy (11%; supplementary information, Table S1). Furthermore, mothers who consumed caffeine were more likely to already have children (49%) compared to mothers who did not consume caffeine during pregnancy (33%; supplementary information, Table S1).

**Table 2.** Overview of demographic information across the individual cohorts.

| Cohort | Country and ancestry | DNA methylation array | N high maternal SEP* (%) | M maternal age (SD) | N Maternal smoking (%)** | N parity > 0 (%) | M BMI | M gestational age in weeks |
| --- | --- | --- | --- | --- | --- | --- | --- | --- |
| **ALSPAC** (N = 729) | United Kingdom; Northern European | 450k | 375 (51%) | 29.79 (4.39) | 77 (11%) | 381 (52.3%) | 22.79 (3.63) | 39.53 (1.52) |
| **BiB** (Asian; N = 353) | United Kingdom; Pakistani | EPIC | 146 (41%) | 28.21 (5.37) | 9 (3%) | 249 (70.5%) | 25.75 (5.23) | 39.17 (1.52) |
| **BiB** (White EUR; N = 306) | United Kingdom; Northern European | EPIC | 125 (41%) | 26.98 (6.15) | 93 (30%) | 159 (52%) | 27.10 (6.48) | 39.29 (1.88) |
| **Generation R** (N = 798) | The Netherlands; Northern European | 450k | 458 (57%) | 30.15 (4.95) | 109 (14%) | 95 (58.6%) | 23.06 (3.64) | 40.20 (1.48) |
| **INMA** (N = 378) | Spain; Southern European | 450k | 277 (73%) | 31.55 (4.07) | 53 (14%) | 161 (42.6) | 23.79 (4.44) | 41.06 (1.34) |
| **EDEN** (N = 162) | France; Southern & Northern European | 450k | 113 (70%) | 31.94 (4.10) | 26 (16%) | 324 (40.6) | 23.52 (4.64) | 39.51 (1.33) |
| **MoBa1** (N = 999) | Norway; Northern European | 450k | 761 (76%) | 29.93 (4.35) | 287 (29%) | 580 (58.1) | 24.02 (4.18) | 39.95 (1.56) |
| **Total or M** (N = 3,725) | - | - | 2,255 (61%) | 30.01(4.60) | 654 (18%) | 1949 (52%) | 23.61 (4.12) | 39.94 (1.51) |

Note. M = Mean. SD = Standard deviation. In the Total row, average caffeine content was calculated by weighting by the inverse variance for each cohort. \* High maternal socio-economic position (SEP): maternal education ≥ high school diploma. \*\* continued smoking during pregnancy. Parity = one or more previous pregnancies. EU = European.

#### Association between maternal caffeine consumption and offspring cord blood DNA methylation

Results of the quality control checks for the individual cohort results can be found in supplementary information, Figures S1 to S15. The overall EWAS meta-analysis results of the caffeine models can be found in Table 3. After adjusting for multiple testing, one CpG site (cg19370043, nearest gene *PRRX1*) was negatively associated with total maternal caffeine consumption (estimate = -2.18x10^-05^; 95% CI: -2.98x10^-05^ to -1.37x10^-05^; *P* = 1.32x10^-07^) and one CpG sites (cg14591243, nearest gene *STAG1*) with caffeine consumed from cola in mg/day (estimate = 2.78x10^-05^, 95% CI: 2.779x10^-05^ to 2.781x10^-05^, *P* = 5.59x10^-09^) (Table 3). Only the total maternal caffeine-associated CpG site survived the leave-one-out analysis. For the cola-associated CpG site, the leave-one-out analysis indicated that that MoBa was driving the effect (supplementary information, Figure S16 and S17).

**Table 3.** A summary of results of each EWAS model from the probe-level analysis

| Model* | CpGs with FDR-corrected P-value <0.05 | CpGs surviving leave-one-out analysis | Meta-analysis sample size | Genomic inflation factor ($\lambda$)** |
| --- | --- | --- | --- | --- |
| **Any vs. no caffeine** |  |  |  |  |
| All offspring (minimally adjusted) * | 0 | n.a. | 3731 | 0.97 |
| All offspring (adjusted for covariates) | 0 | n.a. | 3731 | 0.97 |
| Female offspring (adjusted for covariates) | 0 | n.a. | 1797 | 0.99 |
| Male offspring (adjusted for covariates) | 0 | n.a. | 1934 | 1.00 |
| All offspring (adjusted for covariates and gestational age) | 0 |  | 3731 | 0.97 |
| **Caffeine in mg/day** |  |  |  |  |
| All offspring (minimally adjusted)* | 33 | n.a. | 3731 | 1.03 |
| All offspring (adjusted for covariates) | 1 | 1 (100%) | 3731 | 1.00 |
| Female offspring (adjusted for covariates) | 0 | n.a. | 1797 | 1.00 |
| Male offspring (adjusted for covariates) | 0 | n.a. | 1934 | 1.04 |
| All offspring (adjusted for covariates and gestational age) | 0 |  | 3731 | 0.99 |
| **Caffeine from coffee** |  |  |  |  |
| All offspring (adjusted for covariates) | 0 | n.a. | 2779 | 1.02 |
| **Caffeine from tea** |  |  |  |  |
| All offspring (adjusted for covariates) | 0 | n.a. | 3477 | 1.00 |
| **Caffeine from cola** |  |  |  |  |
| All offspring (adjusted for covariates) | 1 | 0 | 2610 | 1.00 |

*Note. \* only adjusted for estimated cell counts and 20 surrogate variables. Covariates: maternal age, maternal smoking, maternal parity, maternal education, maternal BMI, estimated cell counts and 20 surrogate variables. \*\* The genomic inflation factor ( $\lambda$ ) estimates the extent of bulk inflation of EWAS P-values and the excess false positive rate. 1 = no inflation; > 1 some evidence of inflation.*

#### Beverage-specific effects

Figure 1 displays DNA methylation at the two CpG sites discovered in the probe-level analysis across the different caffeine models. If these associations were truly driven by caffeine exposure, we would expect to see similar associations of these CpG sites across the different beverage models. The association between DNA methylation at cg19370043 and maternal total caffeine consumption appears to be mostly driven by coffee and not by caffeine from tea or cola (Figure 1). For coffee intake specifically, the P-value was < 0.05, but the association with coffee did not survive adjustment for multiple testing in the EWAS meta-analysis, probably because of the lower statistical power to detect small effects in the coffee compared to total caffeine analysis (n total caffeine = 3,731 vs. n coffee = 2,779; because the total caffeine model included mothers with missing beverage-specific data). The association between DNA methylation at cg14591243 appears to be specific to cola consumption rather than general caffeine consumption.

**Figure 1.**
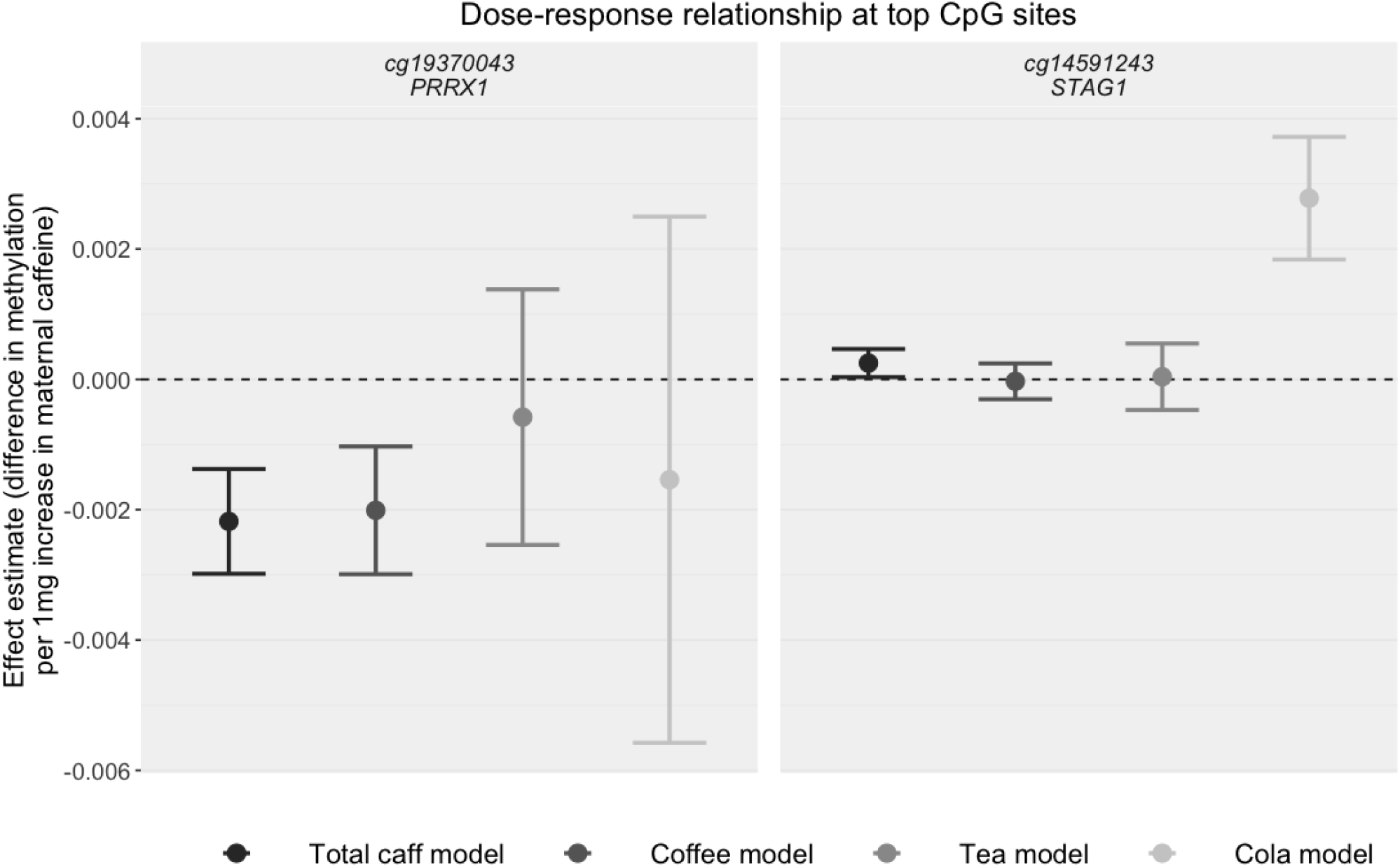
Effect size estimates at the top CpG sites found in the probe-level analysis. Total caff model = total caffeine. Coffee model = caffeine from coffee, Tea model = caffeine from tea, Cola model = caffeine from cola. Error bars represent 95% Confidence Intervals.

### Differentially methylated regions (DMR) meta-analysis

The regional meta-analysis implemented using DMRff detected 22 differentially methylated regions for total maternal caffeine consumption with at least two and a maximum of 15 consecutive CpG sites (*P*_Bonferroni_ < 0.05; supplementary information, Table S2). The strongest evidence was found at a region on chromosome 17, with 7 consecutive CpG sites (chr17: 58499679-58499911; estimate = -3.77 x 10^-05^; SE = 5.02 x 10^-06^; *P*_Bonferroni_ = 1.42 x 10^-10^; supplementary information, Table S2). In the any vs. no maternal caffeine consumption model, there was evidence for 11 DMRs (supplementary information, Table S3). The strongest evidence was found at a region on chromosome 6, with 10 consecutive CpG sites (Chr6:31734147-31734554; estimate = -9.44^-03^; SE = 1.37^-03^; *P*_Bonferroni_ = 1.93^-06^). DMR analyses from the individual sources of caffeine revealed 12 DMRs for caffeine consumed from coffee (supplementary information, Table S4), 18 DMRs for caffeine from tea (supplementary results, Table S5), and 14 DMRs for caffeine consumption from cola (supplementary information, Table S6) during pregnancy. The analyses from the sex-stratified models showed evidence for total maternal caffeine being associated with 12 DMRs in cord blood in female sex offspring (supplementary information, Table S7) and 18 DMRs in male sex offspring (supplementary information, Table S8).

For each pairwise combination of models, we calculated the percentage overlap of CpG sites (or closest gene) by dividing the overlap of CpG sites (or genes) between two models by the sum of the models’ unique CpG sites/genes (e.g., percentage crossover any caffeine and total caffeine models: 7/(167 + 63 - 7) = 0.03 * 100 = 3%). There was very little overlap in CpG sites (range percentage overlap: 0-12%) or annotated genes (% overlapping genes: 0-11%) between the different models (supplementary information, Table S9).

#### Functional analysis of DMRs

Neither the functional categories defined by GO terms nor any of the KEGG pathways showed evidence for enrichment in genes annotated to CpG sites in the caffeine-associated DMRs (all FDR adjusted *P*-values > 0.05). The top 5 KEGG pathways and GO terms with the strongest evidence according to the smallest P-values for each list of CpGs in the caffeine-DMRs are available in supplementary information, Table S10.

## Discussion

### Summary & interpretation of findings

We investigated the association of self-reported maternal caffeine consumption during pregnancy with offspring cord blood DNA methylation using data from six international birth cohorts. For the EWAS meta-analysis, probe-level and regional DMR analyses were applied as hypothesis-free approaches to detect associations between maternal caffeine phenotypes and differential methylation levels in cord blood. We compared caffeine consumption during pregnancy across countries and caffeinated drinks, which reduced the potential for cultural confounding, and analysed the CpG sites of the maternal caffeine associated DMRs for their biological function. Results of these analyses show little converging evidence between the associations of the different sources of caffeine, indicating that the associations that we observed are most likely explained by other factors than caffeine exposure.

The probe-level analysis indicated that differences in DNA methylation at two CpG sites were associated with maternal caffeine intake (one with maternal total caffeine consumption and one with maternal caffeine consumption from cola); both showed small effect estimates. The coefficients from the regression analyses represent the change in offspring cord blood DNA methylation at a given CpG site per 1 mg/day increase in maternal caffeine consumption. Putting these results into real life context, and assuming causality and linearity of effects, if the recommended limit of caffeine consumption during pregnancy were doubled from 200 mg/day to 400 mg/day, this would only be associated with 0.4% reduction in DNA methylation at cg19370043. This effect size is in line with the small effect sizes found in the EWAS meta-analysis of adult personal caffeine consumption on DNA methylation by Karabegović and colleagues (10), where an additional cup of coffee (=57mg) was associated with a 0.2% decrease in peripheral blood DNA methylation at a CpG site near *AHRR*, which would be equivalent to a 0.7% decrease in DNA methylation per 200 mg/day of caffeine (0.2% / 57 mg of caffeine per cup of coffee x 200). These estimated effect sizes appear to be much smaller than the estimated effect of smoking, which is the lifestyle exposure with the strongest effect on DNA methylation discovered to date. Sustained smoking during pregnancy was associated with changes of up to ∼7% decrease in offspring cord-blood DNA methylation at the *AHRR* gene (36). As acknowledged by Karabegović et al, because their smoking adjustment did not include the amount of smoking or duration of smoking, the coffee-associated DNA methylation differences might be explained by residual confounding by smoking (10).

In the regional analyses, we identified 12-22 DMRs for each of the caffeine models. Yet, lack of congruence of associations across models, which was evident in the probe-level and regional analysis, provided evidence for beverage-specific effects instead of the effects being driven by caffeine (which is common to all included beverages).

### Strengths & limitations

This was the first international EWAS meta-analysis investigating associations between offspring DNA methylation and caffeine from coffee, tea, and cola during pregnancy. A major strength of this study is the consideration of the effects of other common sources of caffeine besides coffee. Consumption of the different sources of caffeine might be differentially socially patterned, allowing capturing a larger spectrum of the caffeine consuming population. For instance, British and non-European ethnicities consume more caffeinated tea than coffee (6,47). Further, it is indicated that the main source of caffeine might change during pregnancy, with even habitual coffee drinkers preferring caffeinated tea to coffee during pregnancy (48,49). Last, in contrast to previous research, this study assessed maternal caffeine consumption through mg/day instead of cups per day, which is a useful approach to isolate the effect of caffeine, allow comparison between different caffeinated beverages, and a more fine-tuned assessment of the effects of different caffeine dosages. We also adjusted for maternal smoking, an important potential confounder in analyses of caffeine intake, along with other potential confounding variables.

The findings should be considered in the light of the following limitations. Caffeine assessment in the meta-analysis relied on self-report, which might be underestimated (50) or underreported during pregnancy because of social stigma around maternal health behaviours (51,52). However, this would be more obvious for more recent cohorts and less and less likely for older cohorts, such as ALSPAC, due to awareness of the potential toxicity of caffeine in pregnancy emerging only recently. The questions used to assess caffeine consumption in the cohorts only allowed for rough estimations of maternal caffeine consumption (53), which could reduce power. Generalizability of the results to other populations might be limited by examining only first trimester consumption, the relatively low caffeine intake and the tendency for birth cohorts to enrol more advantaged families(16). Effects of caffeine exposure during pregnancy on offspring cord blood DNA methylation were only assessed at regions available on the 450k array, which only covers around 2% of CpG sites of the entire epigenome (54). Thus, differentially methylated CpGs or regions not covered by the array may have been missed in this study. We only assessed offspring DNA methylation in blood. There is some evidence suggesting that DNA methylation levels in blood might be able to proxy for DNA methylation levels in other tissues, yet we cannot rule out that maternal caffeine during pregnancy might be influencing DNA methylation differentially in other tissue types (55). Finally, this study indicates that, if maternal caffeine consumption influences cord-blood DNA methylation, the effect is likely to be small. Although our meta-analysis maximised sample sizes, even larger sample sizes with more variable levels of caffeine consumption may be required to detect small effects of prenatal caffeine exposure on offspring DNA methylation.

### Future research & conclusion

Future research should aim to use a more accurate assessment of caffeine consumption during pregnancy by considering differing types of coffee, brewing times, and cups sizes and/or assessing biomarkers of caffeine such as plasma concentrations of the caffeine metabolite paraxanthine (56). Triangulation strategies may be applied to disentangle confounded from causal effects of caffeine exposure during pregnancy on offspring DNA methylation. These might include: Further exploring the different confounding structures of various caffeinated beverages and considering individual differences in the maternal metabolism of caffeine. Maternal caffeine metabolism might influence intensity of exposure during pregnancy and might change the effect of caffeine on offspring DNA methylation. For instance, studies could conduct analyses using genetic variants that account for differences in caffeine metabolism (57) and/or consider pre-pregnancy caffeine consumption to account for differences in the tolerance to effects of caffeine during pregnancy (53). Also, more assessments of prenatal paternal caffeine consumption would enable the conduction of negative control analyses to investigate intrauterine effects (58,59), as well as investigating the effects of paternal caffeine consumption prior to pregnancy and its effect on offspring DNA methylation in its own right (59).

In conclusion, results of this large scale EWAS meta-analysis indicate little evidence for a strong effect of maternal caffeine consumption during the second trimester of pregnancy on offspring cord blood DNA methylation.

## Data Availability

Access to all data used in the present study can be requested from the individual cohort studies.

