## supplementary information for "Maternal caffeine consumption during pregnancy and offspring cord blood DNA methylation: a meta-analysis of epigenome-wide association studies"

Table of content

Cohort-specific methods	3

Avon Longitudinal Study of Parents and Children (ALSPAC)	3

Born in Bradford	5

MoBa	7

Generation R	9

INfancia y Medio Ambiente (INMA)	11

EDEN	14

Measurement of maternal caffeine intake during pregnancy	16

Quality checks for probe-level analysis	16

Table S1	Demographic variables of the cohorts stratified by caffeine user status	18

Quality control checks	19

Figure S1	Correlation plot ALSPAC	19

Figure S2	Correlation plot BiB (Asian ethnicity)	19

Figure S3	Correlation plot BiB (White European ethnicity)	20

Figure S4	Correlation plot Generation R	20

Figure S5	Correlation plot EDEN	22

Figure S6	Correlation plot INMA	22

Figure S7	Correlation plot MoBa1	23

Figure S8	ALSPAC QQ-Plots of caffeine models	24

Figure S9	BiB (Asian) QQ-Plots of caffeine models	25

Figure S10	BiB (White European) QQ-Plots of caffeine models	26

Figure S11	Generation R QQ-Plots of caffeine models	27

Figure S12	MoBa1 QQ-Plots of caffeine models	28

Figure S13	EDEN QQ-Plots of caffeine models	29

Figure S14	INMA QQ-Plots of caffeine models	30

Figure S15	Precision plots	31

Figure S16	Leave-one-out plot of the prenatal caffeine-associated CpG site (Cg19370043)	32

Figure S17	Leave-one-out plot of the prenatal cola-associated CpG site (Cg14591243)......	32

Figure S18	Correlation matrix of the meta-analysed caffeine models	33

Differentially methylated regions	34

Table S2	Results from the meta-analysis of differentially methylated regions for total maternal caffeine consumption	34

Table S3	Results from the meta-analysis of differentially methylated regions for any vs. no maternal caffeine consumption	35

Table S4	Results from the meta-analysis of differentially methylated regions for maternal coffee consumption	36

Table S5	Results from the meta-analysis of differentially methylated regions for maternal tea consumption	37

Table S6	Results from the meta-analysis of differentially methylated regions for maternal cola consumption	38

Table S7	Results from the meta-analysis of differentially methylated regions for maternal caffeine consumption stratified by female sex	39

Table S8	Results from the meta-analysis of differentially methylated regions for maternal caffeine consumption stratified by male sex	40

Table S9	Crossover of CpG sites and genes of the DMRs of the different caffeine models…………………………………………………………………………………………………………………………41

Table S10	Top 5 GO terms and KEGG pathways for CpGs in DMRs (BP = biological process; MF = molecular function; CC = cell compartment)	42

Cohort-specific acknowledgments	46

Cohort-specific funding statements	47

### Cohort-specific methods

#### Avon Longitudinal Study of Parents and Children (ALSPAC)

##### Description of the cohort

Ethical approval for the study was obtained from the ALSPAC Ethics and Law Committee and the Local Research Ethics Committees. Informed consent for the use of data collected via questionnaires and clinics was obtained from participants following the recommendations of the ALSPAC Ethics and Law Committee at the time.

Pregnant women resident in Avon, UK with expected dates of delivery between 1st April 1991 and 31st December 1992 were invited to take part in the study. 20,248 pregnancies have been identified as being eligible and the initial number of pregnancies enrolled was 14,541. Of the initial pregnancies, there was a total of 14,676 foetuses, resulting in 14,062 live births and 13,988 children who were alive at 1 year of age. When the oldest children were approximately 7 years of age, an attempt was made to bolster the initial sample with eligible cases who had failed to join the study originally. As a result, when considering variables collected from the age of seven onwards (and potentially abstracted from obstetric notes) there are data available for more than the 14,541 pregnancies mentioned above: The number of new pregnancies not in the initial sample (known as Phase I enrolment) that are currently represented in the released data and reflecting enrolment status at the age of 24 is 906, resulting in an additional 913 children being enrolled (456, 262 and 195 recruited during Phases II, III and IV respectively). The phases of enrolment are described in more detail in the cohort profile paper and its update (see footnote 5 below). The total sample size for analyses using any data collected after the age of seven is therefore 15,447 pregnancies, resulting in 15,658 foetuses. Of these 14,901 children were alive at 1 year of age. Of the original 14,541 initial pregnancies, 338 were from a woman who had already enrolled with a previous pregnancy, meaning 14,203 unique mothers were initially enrolled in the study. As a result of the additional phases of recruitment, a further 630 women who did not enrol originally have provided data since their child was 7 years of age. This provides a total of 14,833 unique women (G0 mothers) enrolled in ALSPAC as of September 2021. G0 partners were invited to complete questionnaires by the mothers at the start of the study and they were not formally enrolled at that time. 12,113 G0 partners have been in contact with the study by providing data and/or formally enrolling when this started in 2010. 3,807 G0 partners are currently enrolled.

Please note that the study website contains details of all the data that is available through a fully searchable data dictionary and variable search tool:

<http://www.bristol.ac.uk/alspac/researchers/our-data/>

##### DNA methylation data

As part of the Accessible Resources for Integrated Epigenomic Studies (ARIES, http://www.ariesepigenomics.org.uk/) project, DNA methylation was generated for 1018 mother-offspring pairs from the ALSPAC cohort, using the Infinium HumanMethylation450 BeadChip array (Illumina Inc., San Diego, United States). ARIES participants were selected based on availability of DNA samples at two time points for the mother (antenatal and at follow-up when the offspring were adolescents) and at three time points for the offspring (neonatal, childhood (age 7), and adolescence (age 17)). The current study used child cord blood at birth and whole blood at age 7. Consent for biological samples has been collected in accordance with the Human Tissue Act (2004).

Methods for methylation measurements in ALSPAC have been described previously (Relton et al., 2015). Briefly, cord blood was collected according to standard procedures. DNA methylation assays and data pre-processing was performed at the University of Bristol as part of the ARIES project. DNA was extracted using standard protocol and was bisulfite-converted using the Zymo EZ DNA MethylationTM kit (Zymo, Irvine, CA). DNA methylation was then measured using the Infinium HM450 BeadChip assay (Illumina Inc, San Diego, CA), according to the standard protocol. Arrays were scanned using an Illumina iScan. An initial review of data quality was assessed using GenomeStudio (version 2011.1). A semi-random approach (sampling criteria were in place to ensure that all time points were represented on each array) was used to distribute ARIES samples across slides to minimize the possibility of potential confounding by batch. Data were normalised using the meffil R package (Min et al, 2018) using the functional normalisation approach.

##### Definition of variables and covariates

- **Maternal coffee consumption:** Coffee intake was initially assessed in cups per weekday and cups per weekend during 18-weeks gestation. Total cups of coffee were derived by adding up the cups of coffee consumed on weekdays and weekends. Cups of coffee were then transformed to mg/day by: (cups/week*57)/7.
- **Maternal tea consumption:** Tea intake was initially assessed in cups per weekday and cups per weekend during 18-weeks gestation. Total cups of tea were derived by adding up the cups of tea consumed on weekdays and weekends. Cups of tea were then transformed to mg/day by: (cups/week*27)/7.
- **Maternal cola consumption:** Cola intake was initially assessed in cans per weekday and weekend during 18-weeks gestation. Total cans of cola were derived by adding up the cans of cola consumed on weekdays and weekends. Cans were then transformed to mg/day by: (cans/week*20)/7.
- **Maternal total caffeine consumption:** Total maternal caffeine intake during pregnancy in mg/day, summing caffeine from tea, coffee and cola drinks. NAs were treated as 0, unless tea, coffee *and* cola were all missing, in which case the variable was coded as missing.
- **Maternal education** (as proxy for maternal socioeconomic position): Maternal education was assessed in week 32 of gestation and coded as an ordinal variable: "Vocational" = 1, “O level” (at 16, equivalent to lower grades of ordinary-level) = 2, "A level" (ordinary-level school-leaving certificate (at 16) = 3, and “Degree” (advanced-level school-leaving certificate (post-16)/degree) = 4.
- **Maternal smoking during pregnancy** was assessed as an ordinary variable representing 0 = no or early smoking during pregnancy, 1 = Stopped before the second trimester of pregnancy and 2 = Smoking in the third trimester or throughout pregnancy.
- **Maternal age** continuous numeric variable in years assessed at birth of study child.
- **Maternal BMI** was assessed continuous numeric variable in years at 12-weeks gestation.
- **Parity** has been assessed at 18-weeks gestation as number of previous pregnancies resulting in either a livebirth or a stillbirth.
- **Gestational age** was calculated (in days) based on the date of the mother’s last menstrual period (LMP) when the mother was certain of this, but for uncertain LMPs and conflicts with clinical assessment the ultrasound assessment was used. Where maternal report and ultrasound assessment conflicted, an experienced obstetrician reviewed clinical records and made a best estimate.
- **Offspring sex** was taken from obstetric records.

#### Born in Bradford

##### Description of cohort

Born in Bradford (BiB; <https://borninbradford.nhs.uk> ) is a longitudinal multi-ethnic birth cohort study aiming to examine the impact of environmental, psychological and genetic factors on maternal and child health and wellbeing (Wright et al., 2013). Bradford is a city in the North of England with high levels of socio-economic deprivation and ethnic diversity. Women were recruited at the Bradford Royal Infirmary at 24-29 weeks gestation. For those consenting, a baseline questionnaire was completed. The full BiB cohort recruited 12,450 women comprising 13,773 pregnancies and 13,858 children between 2007 and 2010. Results of an oral glucose tolerance test (OGTT) and lipid profiles were obtained on the mothers during pregnancy at recruitment (24-29 weeks gestation), and pregnancy serum, plasma and urine samples have been stored. Cord blood samples have been obtained and stored and DNA extraction has been completed on all cord and pregnancy samples. The cohort is broadly characteristic of the city's maternal population. Mean age of the mothers at study recruitment was 27 years old. Researchers are looking at the links between the circumstances of a child's birth, the context in which they grow up, their health and well-being and their educational progress.

For DNAm assays, 1000 mother-child pairs were selected (2000 individuals) from the participants who had completed a pregnancy OGTT (85% of BiB participants completed an OGTT), genome wide data on both mother and offspring (at the time of selection ~65% of those with OGTT) and were of either Pakistani or white British ethnic origin (the two largest homogeneous ethnic groups jointing reflecting 90% of the cohort). Within these criteria 500 Pakistani and 500 white British mother-child pairs were randomly selected. Ethical approval for the data collection was granted by Bradford Research Ethics Committee (Ref 07/H1302/112). On registration with the study, pregnant mothers gave written informed consent for themselves and on behalf of their child.

##### DNA methylation data

A total of 500 ng high molecular weight DNA was bisulfite-converted using the EZ-96 DNA methylation kit (Zymo Research, Orange, CA, USA). DNAm was quantified using Illumina HumanMethylation EPIC Arrays (Illumina, San Diego, CA, USA). During the data generation process, a wide range of batch variables were recorded in a purpose-built laboratory information management system (LIMS). Sample QC and normalization was performed using with meffil9. Samples failing QC (average probe p > 0.01) were excluded from further analysis. As an additional QC step genotype probes were compared with genotype data from the same individual to identify and remove any sample mismatches. Furthermore, samples failed on control probes (bisulfite 1 and bisulfite 2) were also excluded from the analysis. Finally, 864 samples passed QC. Samples were normalized using functional normalization using meffil (Min et al., 2018). Data was normalised using 7 control probe PCs derived from the technical probes informed by meffil scree plots.

##### Definition of variables and covariates

- **Maternal coffee consumption:** Coffee intake was initially assessed in cups of caffeinated filter/cafetiere coffee and cups of instant coffee consumed per day during 26-28 weeks of gestation. Total cups of coffee were derived by adding up the number of cups of filter/cafetiere coffee and cups of instant coffee consumed per day. Missing data were treated as 0, unless caffeinated filter/cafetiere coffee *and* instant coffee were all missing, in which case the variable was coded as missing. Cups of coffee were then transformed to mg/day by: (cups/week*57)/7.
- **Maternal tea consumption:** Tea intake was initially assessed in cups of caffeinated tea per day during 26-28 weeks of gestation. Cups of tea were then transformed to mg/day by: (cups/week*27)/7.
- **Maternal cola consumption:** Cola intake was assessed in cups of regular, caffeinated cola per day and cups of caffeinated diet cola per day. Missing data were treated as 0, unless caffeinated regular cola *and* diet cola were all missing, in which case the variable was coded as missing Cups were then transformed to mg/day by: (cups/week*20)/7.
- **Maternal total caffeine consumption:** Total maternal caffeine intake during pregnancy in mg/day, summing caffeine from tea, coffee and cola drinks. NAs were treated as 0, unless tea, coffee *and* cola were all missing, in which case the variable was coded as missing.
- **Maternal education** (as proxy for maternal socioeconomic position): Maternal education was assessed in week 26-28 weeks of gestation and coded as an ordinal variable: " <5 GCSE equivalent " = 1, “5 GCSE equivalent” (at 16, equivalent to lower grades of ordinary-level) = 2, " A-level equivalent " (ordinary-level school-leaving certificate (at 16) = 3, and “Higher than A-level” (advanced-level school-leaving certificate (post-16)/degree) = 4.
- **Maternal smoking during pregnancy** was assessed as an ordinary variable representing 0 = no or early smoking during pregnancy, 1 = Stopped before the second trimester of pregnancy and 2 = Smoking in the third trimester or throughout pregnancy.
- **Maternal age** continuous numeric variable in years assessed at 26-28 weeks of gestation.
- **Maternal BMI** was assessed continuous numeric variable in years at 26-28 weeks of gestation**.**
- **Parity** has been assessed at routine healthcare as an integer value.
- **Gestational age** age at completion of questionnaire at weeks 26-28 of gestation (weeks and days).
- **Offspring sex** was assessed at routine healthcare.

#### MoBa

##### Description of the cohort

Participants represent three subsets of mother-offspring pairs from the nation-wide Norwegian Mother, Father and Child Cohort Study (MoBa) (Magnus et al. 2016; <https://www.fhi.no/en/studies/moba/>). MoBa is a prospective population-based pregnancy cohort study conducted by the Norwegian Institute of Public Health. The years of birth for MoBa participants ranged from 1999-2008. MoBa mothers provided written informed consent. MoBa1 is a subset of a larger study within MoBa that included a cohort random sample and cases of asthma at age three years (Haberg et al., 2011). We previously reported an association between maternal smoking during pregnancy and differential DNA methylation in MoBa1 newborns (Joubert et al., 2012). Years of birth were 2002-2004 for children in MoBa1. The establishment of MoBa and initial data collection was based on a license from the Norwegian Data Protection Agency and approval from The Regional Committees for Medical and Health Research Ethics. MoBa is currently regulated by the Norwegian Health Registry Act. The study was approved by the Regional Committee for Ethics in Medical Research, Norway (#2017/1362). In addition, MoBa1 was approved by the Institutional Review Board of the National Institute of Environmental Health Sciences, USA. The consent given by the participants does not allow for storage of data on an individual level in repositories or journals. Researchers who want access to data sets for replication should submit an application to <http://www.helsedata.no/>. Access to data sets requires approval from The Regional Committee for Medical Research Ethics in Norway and an agreement with MoBa.

##### DNA methylation data

Details of the DNA methylation measurements and quality control for the MoBa1 participants were previously described and the same protocol was implemented for the MoBa2 participants (Joubert et al., 2012). Briefly, umbilical cord blood samples were collected and frozen at birth at -80°C. All biological material was obtained from the Biobank of the MoBa study (Paltiel et al., 2014). Bisulfite conversion was performed using the EZ-96 DNA Methylation kit (Zymo Research Corporation, Irvine, CA) and DNA methylation was measured at 485,577 CpGs in cord blood using Illumina’s Infinium HumanMethylation450 BeadChip. Raw intensity (.idat) files were handled in R using the minfi package to calculate the methylation level at each CpG as the beta-value (β=intensity of the methylated allele (M)/(intensity of the unmethylated allele (U) + intensity of the methylated allele (M) + 100)) and the data was exported for quality control and processing. Control probes (N=65) and probes on X (N=11 230) and Y (N=416) chromosomes were excluded in both datasets. Remaining CpGs missing > 10% of methylation data were also removed (N=20). Samples indicated by Illumina to have failed or have an average detection p value across all probes < 0.05 (N=49) and samples with gender mismatch (N=13 MoBa1) were also removed. We accounted for the two different probe designs by applying the intra-array normalization strategy Beta Mixture Quantile dilation (BMIQ). After quality control exclusions, the sample sizes were 1,068.

##### Definition of variables and covariates

- **Maternal coffee consumption:** In MoBa coffee intake was assessed based on type of coffee (instant/espresso coffee) and brewing method (boiled/percolated/filtered) at the 17th week of gestation. Cups were transformed to mg of caffeine based on assuming 85 mg of caffeine per cup of boiled/ percolated/ filtered coffee and 60 mg per cup of instant/espresso coffee
- **Maternal tea consumption**: Maternal cups of tea at 17 weeks gestation were transformed to mg based on the assumption that one cup of tea contains 50mg of caffeine
- **Maternal cola consumption:** Cola consumption was measured as consumption of regular and diet Coca Cola/Pepsi in mugs and the transformed to cups (one mug = two cups; one small bottle = four cups; one large bottle, 1.5L = 12 cups). Cups were then transformed based on the assumption that one cup of cola contains 30mg of caffeine
- **Maternal education** was grouped into 4 levels, 0 = not completed high school, 1 = High school, 2= some college and 4 = four or more years of college/university.
- **Maternal smoking during pregnancy** categorical as 0/1, with 1 for any smoking during pregnancy, and 0 for no-smoking during pregnancy.
- **Maternal pre-pregnancy BMI** self-reported continuous numeric variable in kg/m2 at around 16 weeks gestation**.**
- **Gestational age** was calculated (in days) based on the ultrasound measurements taken at first check up in pregnancy (around 16 weeks, at enrolment in the cohort). If ultrasound measurement was not available, this was based on the last menstrual period.
- **Maternal age** continuous numeric variable in years assessed by the Norwegian Medical Birth Registry
- **Parity** was assessed by the Norwegian Medical Birth Registry as the number of previous live births
- **Offspring sex** was provided by the Norwegian Medical Birth Registry.

#### Generation R

##### Description of the cohort

The Generation R Study (<https://generationr.nl/researchers/>) is a population-based prospective cohort study from fetal life onwards in Rotterdam, the Netherlands. A detailed description can be found elsewhere [Kooijman et al, Eur J Epidemiol 2016; Kruithof et al, Eur J Epidemiol 2014]. Briefly, 9,778 pregnant women with a delivery date between April 2002 and January 2006 were included (response rate at baseline 61%). There is ongoing follow-up. The study was approved by Medical Ethical Committee of Erasmus MC, University Medical Center Rotterdam and written consent was obtained for all participants. For this analysis, we used singleton live births with epigenome-wide association arrays and information on paternal BMI and complete covariates, giving total sample sizes of 947 at birth and 335 in childhood.

##### DNA methylation data

DNA, 500 ng per sample, extracted (using the salting-out method) from blood samples taken at birth (cord blood) or at the 5-year follow-up underwent bisulfite conversion using the EZ-96 DNA Methylation kit (Shallow) (Zymo Research Corporation, Irvine, USA). Samples were plated onto 96-well plates in no specific order. Samples were processed with the Illumina Infinium HumanMethylation450 BeadChip (Illumina Inc., San Diego, USA), which analyses methylation at 485,577 CpG sites. Preparation and normalization of the HumanMethylation450 BeadChip array data was performed according to the CPACOR workflow [Lehne et al. 2015] using the package minfi in R [R Core Team, 2013]. Probes that had a detection p-value above background (based on sum of methylated and unmethylated intensity values) ≥ 1E-16 were set to missing per array. Next, the intensity values were stratified by autosomal and non-autosomal probes and quantile normalized for each of the six probe type categories separately: type II red/green, type I methylated red/green and type I unmethylated red/green. Beta values were calculated as proportion of methylated intensity value on the sum of methylated+unmethylated+100 intensities. Arrays with observed technical problems such as failed bisulfite conversion, hybridization or extension, as well as arrays with a mismatch between sex of the proband and sex determined by the chr X and Y probe intensities were removed from subsequent analyses. Additionally, only arrays with a call rate > 95% per sample were processed further.

##### Definition of variables and covariates

- **Maternal coffee consumption:** Information about maternal coffee intake was assessed by questionnaire at 18-25 weeks gestational. Pregnant women, who indicated to consume coffee, were asked whether they consumed caffeinated or decaffeinated coffee, both, or other. The next question asked about their average number of cups of per day. As the quantity of cups did not differentiate between caffeinated or decaffeinated coffee intake, mothers who indicated to drink both caffeinated and decaffeinated coffee, needed to be excluded (and so were mothers who indicated to drink other types of coffee than caffeinated or decaffeinated). For mothers who reported to drink decaffeinated coffee, the number of cups of coffee per day was set to zero. To calculate the total caffeine intake from coffee, cups were transformed to mg assuming 57 mg of caffeine per cup.
- **Maternal tea consumption:** Women who indicated to consume tea at 18-25 weeks gestational were asked about whether they consumed caffeinated or decaffeinated tea, both, or other. The next question asked about their average number of cups of per day. As the quantity of cups did not differentiate between caffeinated and decaffeinated tea intake, mothers who indicated to drink both caffeinated and decaffeinated tea, needed to be excluded (and so were mothers who indicated to drink other tea than caffeinated or decaffeinated). For mothers who reported to drink decaffeinated tea (herbal or green tea), the number of cups of tea per day was set to zero. To calculate the total caffeine intake from tea, cups were transformed to mg assuming 27 mg of caffeine per cup.
- **Maternal cola consumption:** Not available.
- **Maternal total caffeine consumption:** To calculate the total caffeine intake, mg/day of coffee and tea were summed up. For this analysis women with missing data on both coffee and tea were excluded. NAs were treated as 0, unless tea *and* coffee were missing, in which case the variable was treated as NA.
- **Maternal education** (as proxy for maternal socioeconomic position): Maternal education was assessed via a questionnaire sent out during pregnancy (after enrolment) and coded into three categories: ‘no education or primary education’, ‘secondary education/high school’, ‘higher education (college or university)’
- **Maternal smoking during pregnancy** was assessed via questionnaires, sent out during each trimester of pregnancy and coded as three categories: ‘no smoking in pregnancy/quit smoking until pregnancy was known (i.e., first trimester only)’ or ‘continued smoking throughout pregnancy’
- **Maternal age** continuous numeric variable in years assessed via a questionnaire sent out during pregnancy
- **Maternal BMI** was calculated from measured height during a visit to the research center during the first trimester of pregnancy and self-reported pre-pregnancy weight assessed via a questionnaire sent out during pregnancy
- **Parity** was assessed via a questionnaire sent out during pregnancy
- **Gestational age** was calculated (in days) based on the date of the mother’s last menstrual period (LMP) when the mother was certain of this and if she had a regular menstrual cycle of 28±4 days. For uncertain LMP and/or irregular cycle, ultrasound assessment was used.
- **Offspring sex** was recorded by midwife/hospital records.

#### INfancia y Medio Ambiente (INMA)

##### Description of cohort

Population-based birth cohorts were established as part of the INMA – INfancia y Medio Ambiente [Environment and Childhood] Project in several regions of Spain following a common protocol (<http://www.proyectoinma.org/> ). This project aims to study the associations between pre- and postnatal environmental exposures and growth, health, and development from early foetal life until adolescence and has been described previously in detail (Guxens et al., 2012). Pregnant women were enrolled during the 1st trimester of pregnancy at public primary health care centers or public hospitals. Detailed measurements were performed using ultrasound and physical examinations and biological samples were collected. Informed consent was obtained from all participants and the study was approved by the Hospital Ethics Committees in each participating region. Blood DNA methylation data was assessed with the Infinium HumanMethylation450 BeadChip in INMA Sabadell subcohort, at birth, 4y and 9y. In the current study we participated with methylation data assessed at birth. Data is available for 385 out of the 742 children enrolled in the subcohort. We selected those children with good DNA quality, with follow-up information, with genetic data (when possible) and mostly of European ancestry.

##### DNA methylation data

Cord blood was extracted using the Chemagen kit (Perkin Elmer). DNA concentration was determined by a NanoDrop spectrophotometer (Thermo Scientific) and with the Quant-iT PicoGreen dsDNA Assay Kit (Life Technologies). Blood methylation data was produced in two laboratories: the Genome Analysis Facility of the University Medical Center Groningen (UMCG) in Holland as part of the MeDALL project, and the Bellvitge Biomedical Research Institute (IDIBELL) in Barcelona as part of the BREATHE project. Both laboratories randomized the samples in batches and followed the Illumina protocol for the Infinium HumanMethylation450 BeadChip. Briefly, 500 ng of DNA was bisulfite-converted using the EZ 96-DNA methylation kit, and DNA methylation was measured through hybridization on the BeadChips. BeadChips were scanned with an Illumina iScan and image data was uploaded into the Methylation Module of Illumina’s analysis software GenomeStudio, and converted in β-values. Two blood samples with overall low quality (MethylAid package) (van Iterson et al. 2014), and three blood samples discordant for sex (shinyMethyl package) (Fortin et al., 2014) were removed. After applying a stringent detection p-value of 1.10E-16 (Lehne et al. 2015), 18 blood samples with a call rate <98% were excluded. 7,136 probes with a call rate <95%. Control probes, probes with cross-hybridizing problems, and probes designed to detect genetic polymorphisms were flagged. Data was normalized with the functional normalization method with prior background correction with Noob implemented in the minfi package (Aryee et al. 2014).

##### Definition of variables and covariates

- **Maternal coffee consumption:** Coffee intake was assessed in cups per day at week 12 of pregnancy (and it referred to the intake from the last menstruation to the first prenatal visit (10-13 weeks of pregnancy). A semi-quantitative validated FFQ of 101 items (Vioque et al., 2013) was used with 9 possible responses from “never or less than once per month” to “six or more per day”. Based on these responses daily consumption of coffee was calculated. Cups of coffee were then transformed to mg/day by: cups/day*57.
- **Maternal tea consumption:** Tea and herbal infusion intake was assessed in cups per day at week 12 of pregnancy in the same way as coffee consumption. Cups of tea were then transformed to mg/day assuming 27 mg of caffeine per cup.
- **Maternal cola consumption:** Regular and light soda intake was assessed in glasses per day at week 12 of pregnancy. Glasses of soda were then transformed to mg/day assuming 20mg of caffeine per glass of cola.
- **Maternal total caffeine consumption:** Total maternal caffeine intake during pregnancy in mg/day, summing caffeine from tea, coffee and cola drinks.
- **Maternal total caffeine consumption:** Total maternal caffeine intake during pregnancy in mg/day, summing caffeine from tea, coffee and cola drinks.
- **Maternal education** (as proxy for maternal socioeconomic position): Maternal education was assessed at week 12 of gestation and coded as an ordinal variable: 1 = without studies/primary studies unfinished, 2 = primary studies, 3 = secondary, and 4 = University.
- **Maternal smoking during pregnancy** was assessed as an ordinary variable representing 0 = no or quit smoking before second trimester, 1 = Smoking in the third trimester or throughout pregnancy.
- **Maternal age** continuous numeric variable in years assessed at enrolment.
- **Maternal BMI** Maternal pre-pregnancy BMI was calculated from measured height and self-reported pre-pregnancy weight collected using a questionnaire at enrolment (week 12 of pregnancy). Reported pre-pregnancy weight was highly correlated with measured weight at 12 weeks of pregnancy in INMA (r = 0.96; P < 0.0001).
- **Parity** based on previous born children (previous stillbirths included, abortions excluded), asked at 12 weeks assessment. It was coded as 0 = no previous pregnancy and 1 = one or more previous pregnancies.
- **Gestational age** was calculated (in weeks) based on the date of the mother’s last menstrual period when the mother was certain of this, corrected with the information from the ultrasound assessment. It was transformed to days by multiplying it by 7.
- **Offspring sex** was taken from obstetric records.

Infinium DNA methylation microarrays. Bioinformatics. 15:30(10).

Bakulski et al., 2016, DNA methylation of cord blood cell types: applications for mixed cell birth studies. Epigenetics. 11(5):354-62.

Casas M, Chatzi L, Carsin A-E, Amiano P, Guxens M, Kogevinas M, et al. Maternal pre-pregnancy overweight and obesity, and child neuropsychological development: two Southern European birth cohort studies. Int. J. Epidemiol. 2013;42:506–17.

Fortin et al., 2014, Functional normalization of 450k methylation array data improves replication in large cancer studies. Genome Biol. 15(12):503.

Guxens, M., Ballester, F., Espada, M., Fernandez, M.F., Grimalt, J.O., Ibarluzea, J., Olea, N., Rebagliato, M., Tardon, A., Torrent, M. et al. (2012) Cohort Profile: the INMA--INfancia y Medio Ambiente--(Environment and Childhood) Project. Int. J. Epidemiol., 41, 930-940.

Houseman EA, Accomando WP, Koestler DC, Christensen BC, Marsit CJ, Nelson HH, et al. DNA methylation arrays as surrogate measures of cell mixture distribution. BMC Bioinformatics. 2012;13:86

Lehne et al., 2015. A coherent approach for analysis of the Illumina HumanMethylation450 BeadChip improves data quality and performance in epigenome-wide association studies. Genome Biology. 15; 16:37

Reinius LE, Acevedo N, Joerink M, Pershagen G, Dahlén SE, Greco D, Söderhäll C, Scheynius A, Kere J.[Differential DNA methylation in purified human blood cells: implications for cell lineage and studies on disease susceptibility.](http://www.ncbi.nlm.nih.gov/pubmed/22848472)PLoS One. 2012;7(7):e41361. doi: 10.1371/journal.pone.0041361.

Van Iterson M, et al. 2014, MethylAid : visual and interactive quality control of large Illumina 450k datasets, Bioinformatics, 30(23):3435-7.

Vioque J, Navarrete-Muñoz EM, Gimenez-Monzó D, García-de-la-Hera M, Granado F, Young IS, Ramón R, Ballester F, Murcia M, Rebagliato M, Iñiguez C; INMA-Valencia Cohort Study. Reproducibility and validity of a food frequency questionnaire among pregnant women in a Mediterranean area. Nutr J. 2013 Feb 19;12:26.

#### EDEN

##### Description of cohort

EDEN (study on the pre- and early postnatal determinants of child health and development) (Heude et al., 2016) started recruiting pregnant women in 2003 through two University Hospitals in France (Nancy and Poitiers) with a maternity unit. Mother-child pairs have been followed-up for up to 8-years post-pregnancy. Ethics approval for data collection was granted by the ethics committee of Kremlin Bicêtre and the Commission Nationale Informatique et Liberté.

Women were accounted as eligible for the study if they attended the maternity unit before week 24 of amenorrhoea. Further eligibility criteria were a singleton pregnancy, no diagnosed diabetes before pregnancy, French literacy and not planning to move outside of the recruitment area within the next 3 years. Out of 3,758 eligible women that were recruited between 2003 and 2006, 1,034 women enrolled in Nancy and 968 in Poitiers, resulting in a total sample size of 2,002 pregnant mothers (response rate = 53%). Data about maternal and offspring data was collected at 3 clinic visits for mothers, (time-points: 24-28 weeks of amenorrhea, delivery, 5-6 years post-pregnancy), and 4 clinic visits for offspring (time-points: birth, 1-, 3-, 6-years of age). Furthermore, data was collected through one prenatal questionnaire (24-28 week amenorrhoea) and 8 postnatal questionnaires (time-points: 4-months; 8-months; 1-, 2-, 3-, 4-, 5- and 8-years postnatally).

##### DNA methylation data

For the DNA methylation subsample a random sample of 150 children was selected amongst children who met the following requirements: (1) Participated in the five-year follow-up assessment, (2) had consent for the collection of cord blood and peripheral blood at the age of five, and (3) had a DNA methylation sample with sufficient quality. Data was assessed using the 450k microarray and after quality control checks, 439,306 CpG sites were available for analysis (Merid et al., 2020). Estimated cell proportion types were estimated using the R-package minfi (Aryee et al., 2014). Principal components analysis (PCA) was conducted in EDEN and 5 PC were used to control for batch effects (Merid et al., 2020).

##### Definition of variables and covariates

- **Maternal coffee consumption:** Coffee intake was initially assessed using a questionnaire administered during the week 24-28 of gestation, where mothers reported the daily consumption in the first trimester of pregnancy. Total cups of coffee were derived by adding up the cups of coffee consumed per day at home and outdoors. Cups of coffee were then transformed to mg/day by the formula: cups/week*57.
- **Maternal tea consumption:** Tea intake was initially assessed using a questionnaire administered during the week 24-28 of gestation, where mothers reported the daily consumption in the first trimester of pregnancy. Total cups of tea were derived by adding up the cups of tea consumed per day at home and outdoors. Cups of tea were then transformed to mg/day by the formula: cups/week*27.
- **Maternal cola consumption:** regular and light cola intakes were initially assessed using a food frequency questionnaire administered after delivery about the dietary habits in the last trimester of pregnancy. Total volume of cola consumption per day was estimated by multiplying the frequency of consumption by the volume of the most used glass for cola drinking. Total cups of cola were derived by adding up the volumes of regular and light cola consumption per day, divided by 250mL. Cups were then transformed to mg/day by the formula: cups*20.
- **Maternal total caffeine consumption:** Total maternal caffeine intake during pregnancy in mg/day, summing caffeine from tea, coffee and cola drinks. NAs were treated as 0, unless tea, coffee *and* cola were all missing, in which case the variable was coded as missing.
- **Maternal education** (as proxy for maternal socioeconomic position): Highest degree obtained by the mother was assessed at week 24-28 of gestation, as self-reported by the mother, and coded as an ordinal variable: "Vocational school" = 0, “French *baccalauréat* (BAC) degree” = 1, "BAC + 2 years of College" = 2, and “University or higher degree” = 3.
- **Maternal smoking during pregnancy** was assessed as an ordinary variable representing 0 = no or early smoking during pregnancy, 1 = Smoking throughout pregnancy.
- **Maternal age** continuous numeric variable in years assessed at birth of study child
- **Maternal BMI** was assessed continuous numeric variable in kg/m^2^, dividing the weight before the beginning of the pregnancy, as self-reported by the mother at the clinical interview during pregnancy, and squared height in meters, as measured in clinic during pregnancy.
- **Parity** coded as 0 = nulliparous, 1 = with at least one living child.
- **Gestational age** was included as continuous variable (in weeks) based on the date of the mother’s last menstrual period (LMP) and the date of birth of the child.
- **Offspring sex** was taken from obstetric records.

#### Measurement of maternal caffeine intake during pregnancy

A continuous total caffeine score was calculated by summing the caffeine content from each caffeinated drink in mg/day and allowing for partially missing data by only excluding participants if they had missing data on all three caffeine variables (e.g., if a mother reported to consume two cups of tea per day but had missing data for cups of coffee and cola, she would be given a total caffeine score of 2 x 27 = 54 mg/day of caffeine). Further, to control for data entry errors, participants were excluded if they reported consuming more than 5 standard deviations from the mean (equivalent to roughly 5 x 82mg/day = 410 mg/day caffeine, which is approx. 7 cups of coffee or 15 cups of tea). Quality checks for the cohort specific analysis results included correlation matrices of the beta coefficients for each of the models. Also, the distributions of the P-values in QQ-plots were plotted and Lambda values were calculated. Further, precision plots were generated by plotting 1/median standard error against the square root of the sample size of each cohort.

To find out which gene pathways are linked to the CpG sites of the caffeine- associated DMRs, a gene ontology analysis was run using the R package *missMethyl* (38). Due to the unequal number of CpG sites per gene assessed on the 450k array, some genes are overrepresented and more likely to show up as enriched in gene set analyses (Geeleher et al., 2013). *missMethyl* accounts for this bias by considering the probability of a gene pathway being selected in accordance with the number of probes per gene on the array.

#### Quality checks for probe-level analysis

As expected, most of the models using the different caffeine phenotypes correlated moderately or highly. Only the model using caffeine from cola showed consistently very low correlations with the other model coefficients. Furthermore, the models using caffeine from tea and cola showed low and sometimes even negative correlations. This could be due to few mothers consuming caffeine from cola or indicate a differential effect of cola, tea, and coffee on DNA methylation, potentially through a different substance than caffeine or a different confounding structure. Lastly, the sex stratified models showed low correlations, most likely due to the small sample size in these models. Alternatively, this might indicate a differential effect of caffeine for male and female sex offspring. Visual inspection of the QQ-plots and Lambda values ranging from 0.99-1.07 indicated that most of the P-values of the models are randomly distributed (Figures S8 to S14). As expected, the precision plots (Figure S15) showed that the cohorts with lower sample sizes had less precise estimates. MoBa1 showed most precise estimates throughout all models, except for the tea models, where Generation R showed the most precise estimates. Furthermore, the model using caffeine from cola as the exposure variable were found to be less precise in all cohorts except for MoBa1, most likely because of low consumption of caffeine from cola in the other cohorts. ALSPAC showed relatively low precision in the any vs. no caffeine and coffee models, most likely because of few mothers abstaining from caffeine during pregnancy and mothers consuming more caffeine from tea than coffee. Correlation estimates were highest for coffee and lowest for cola (range correlation estimates: Coffee = 0.70-0.75; tea = 0.38-0.41; cola = 0.15-0.17; Figures S1 to S8). This could indicate that cola consumption in our sample was higher confounded than consumption of the other caffeinated beverages.

#### Table S1 Demographic variables of the cohorts stratified by caffeine user status

| **Cohort** | **Users** | | | | | | | **Non-users** | | | | | | |
| --- | --- | --- | --- | --- | --- | --- | --- | --- | --- | --- | --- | --- | --- | --- |
|  | **N (%)** | **M age (SD)** | **N any smoking preg.(%)** | **N high SEP(%)** | **N parity (%)** | **M BMI**  **(SD)** | **M gest. age (SD)** | **N (%)** | **M age (SD)** | **N any smoking preg.(%)** | **N High SEP(%)** | **N parity (%)** | **M BMI (SD)** | **M gest. age(SD)** |
| **ALSPAC** | 664 (91%) | 29.71 (4.38) | 72 (11%) | 334 (50%) | 348 (52%) | 22.81 (3.61) | 39.52 (1.52) | 65 (9%) | 30.68 (4.43) | 5 (8%) | 41 (61%) | 33  (50.8%) | 22.48 (3.79) | 39.65 (1.43) |
| **BiB**  **(Asian)** | 284 (80%) | 28.43 (5.11) | 8 (3%) | 117 (41%) | 75 (26%) | 25.77 (5.17) | 26.61 (2.40) | 69 (20%) | 27.30 (6.32) | 1 (1%) | 29 (42%) | 29 (42%) | 25.70 (5.51) | 26.64 (1.88) |
| **BiB**  **(White EU)** | 262 (86%) | 26.88 (6.24) | 86* (33%) | 104 (40%) | 139 (53%) | 27.25 (6.57) | 26.62 (1.94) | 44 (14%) | 27.55 (5.64) | 7* (16%) | 21 (48%) | 20 (46%) | 26.22 (5.96) | 26.51 (1.48) |
| **Generation R** | 650 (81%) | 32.00 (4.20) | 101* (16%) | 448 (70%) | 277* (42.6) | 23.13 (3.70) | 40.19 (1.51) | 148 (19%) | 31.65 (3.62) | 8 *(5%) | 106 (72%) | 47* (32%) | 22.73 (3.34) | 40.21 (1.41) |
| **INMA** | 326 (86%) | 31.52 (4.02) | 48 (15%) | 241 (74%) | 140 (42.9) | 23.68 (4.19) | 39.77 (1.36) | 52 (14%) | 31.73 (4.39) | 5 (10%) | 36 (69%) | 21 (40%) | 24.49 (5.77) | 39.83 (1.60) |
| **EDEN** | 144 (89%) | 30.07 (5.04) | 25 (17%) | 82 (57%) | 83 (57.6) | 23.73 (4.76) | 39.55 (1.36) | 18 (11%) | 30.78 (4.22) | 1 (6%) | 11 (61%) | 12 (67%) | 30.78 (4.22) | 39.17 (1.10) |
| **MoBa1** | 757  (76%) | 30.19* (4.22) | 241* (32%) | 576 (76%) | 461* (61%) | 24.13 (4.32) | 39.93 (1.60) | 242  (24%) | 29.12* (4.63) | 46* (19%) | 186 (77%) | 49* (20%) | 23.69 (3.67) | 40.00 (1.40) |
| **Total or M** | 3,087 (83%) | 30.41 (4.43) | 581 (20%) | 1,902 (62%) | 1,523 (49%) | 23.64 (4.13) | 38.57 (1.58) | 638 (17%) | 30.26 (4.42) | 73 (11%) | 430 (67%) | 211 (33%) | 23.64 (3.90) | 38.21 (1.45) |

*Note. M = Mean. SD = Standard deviation. In the Total row, average caffeine content was calculated by weighting by the inverse variance for each cohort. Mothers were categorised as non-user of caffeine if they indicated to consume zero mg/day of coffee, tea, or cola. * significant difference between users and non-users according to P-value < 0.05. High maternal socio-economic position (SEP): maternal education >= continued education after high school. Gestational age in BiB was assessed between 26-28 weeks of gestation.* *Parity = one or more previous pregnancies. EU = European. preg. = pregnancy. gest. = gestational age.*

### Quality control checks

#### Figure S1 Correlation plot ALSPAC

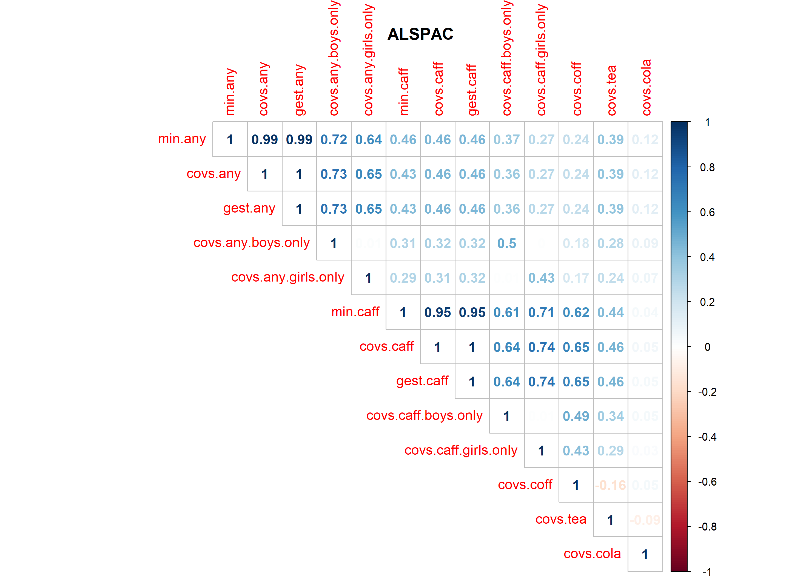

#### Figure S2 Correlation plot BiB (Asian ethnicity)

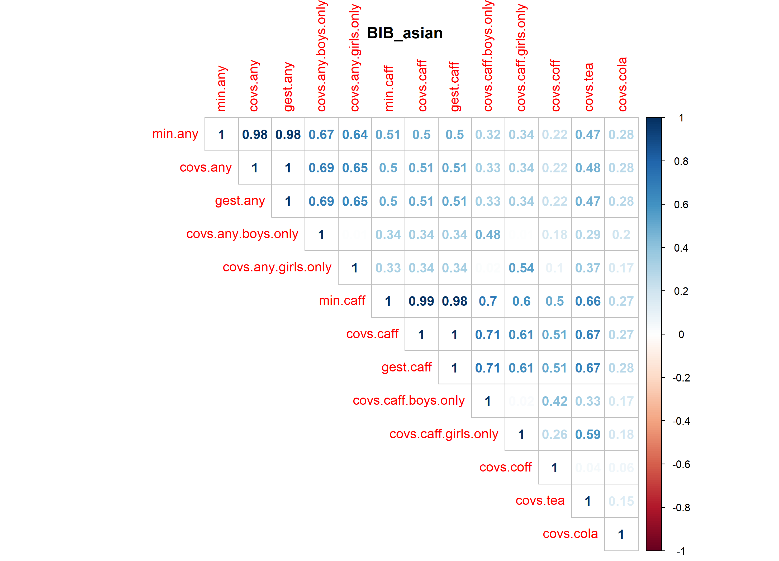

#### Figure S3 Correlation plot BiB (White European ethnicity)

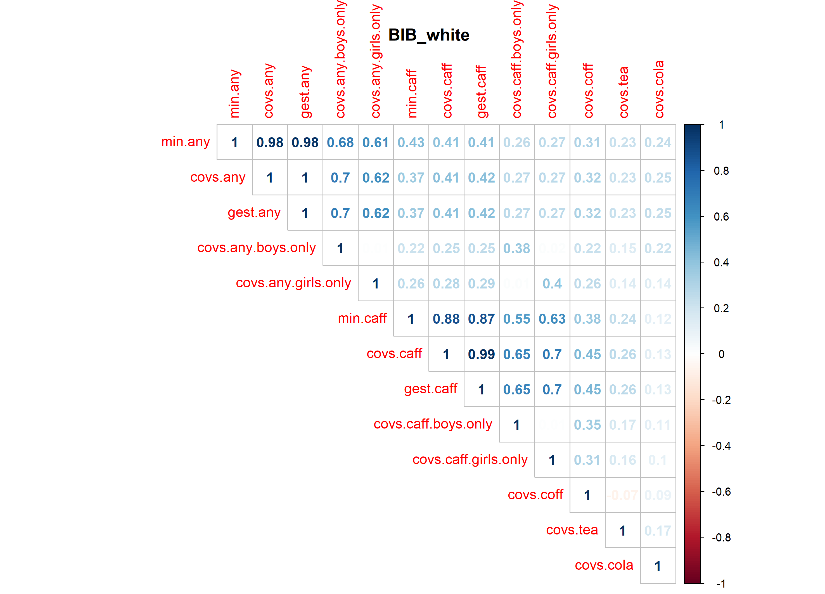

#### Figure S4 Correlation plot Generation R

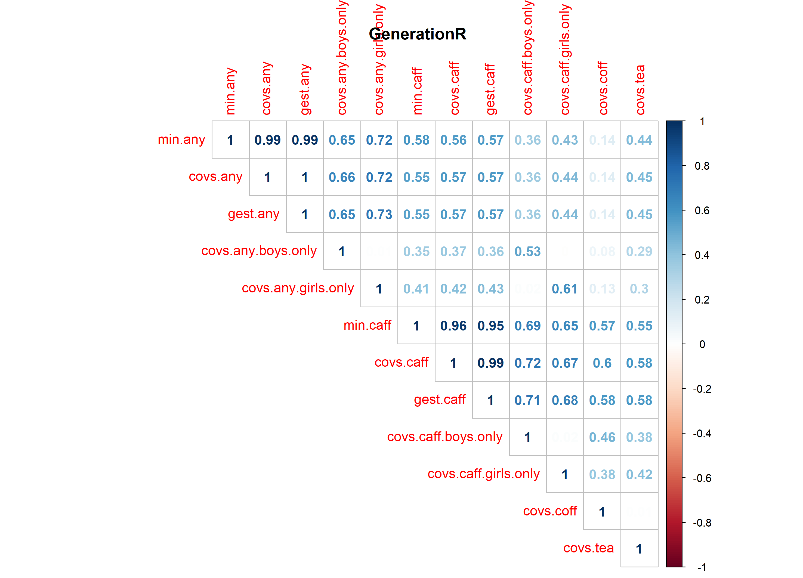

#### Figure S5 Correlation plot EDEN

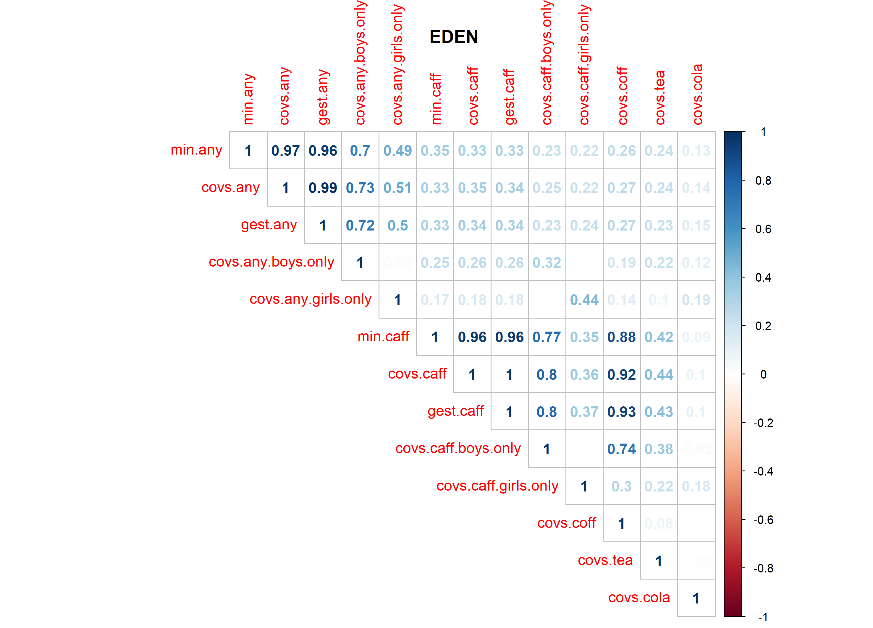

#### Figure S6 Correlation plot INMA

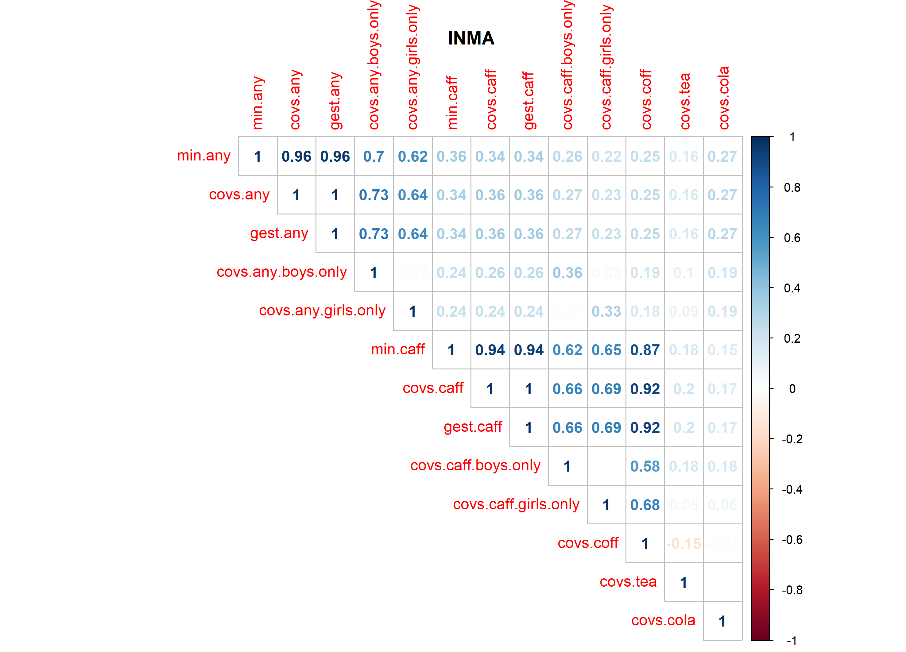

#### Figure S7 Correlation plot MoBa1

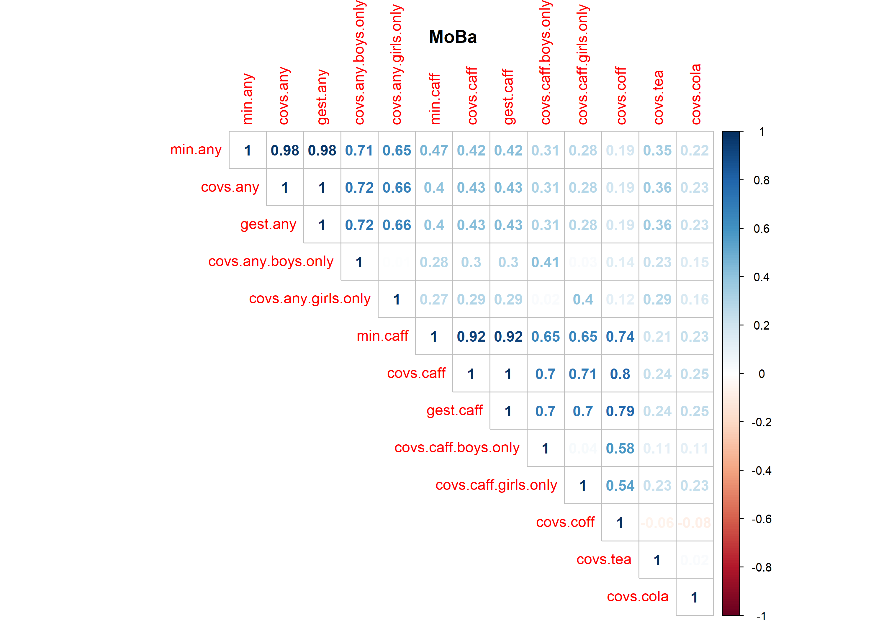

#### Figure S8 ALSPAC QQ-Plots of caffeine models

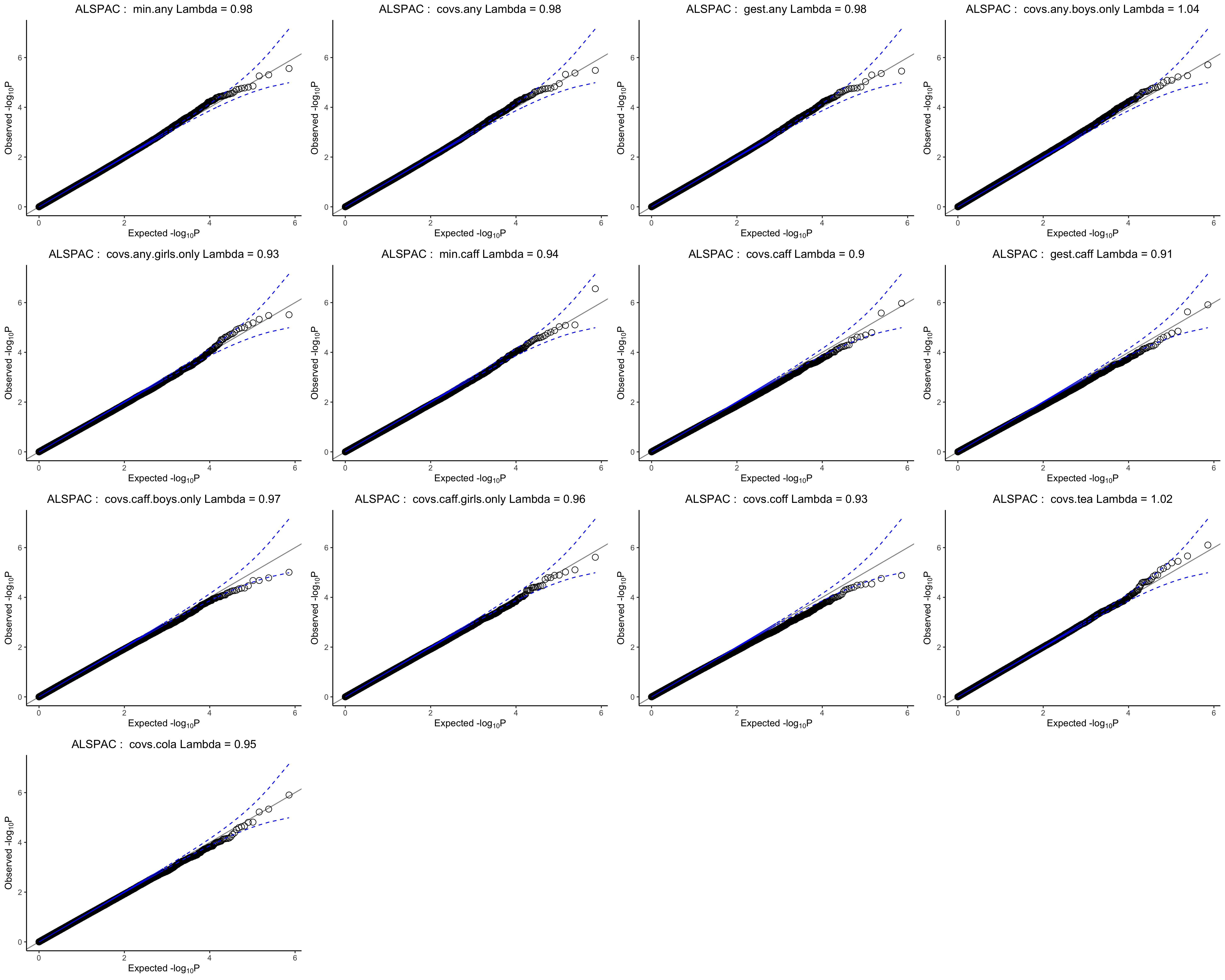

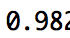

#### Figure S9 BiB (Asian) QQ-Plots of caffeine models

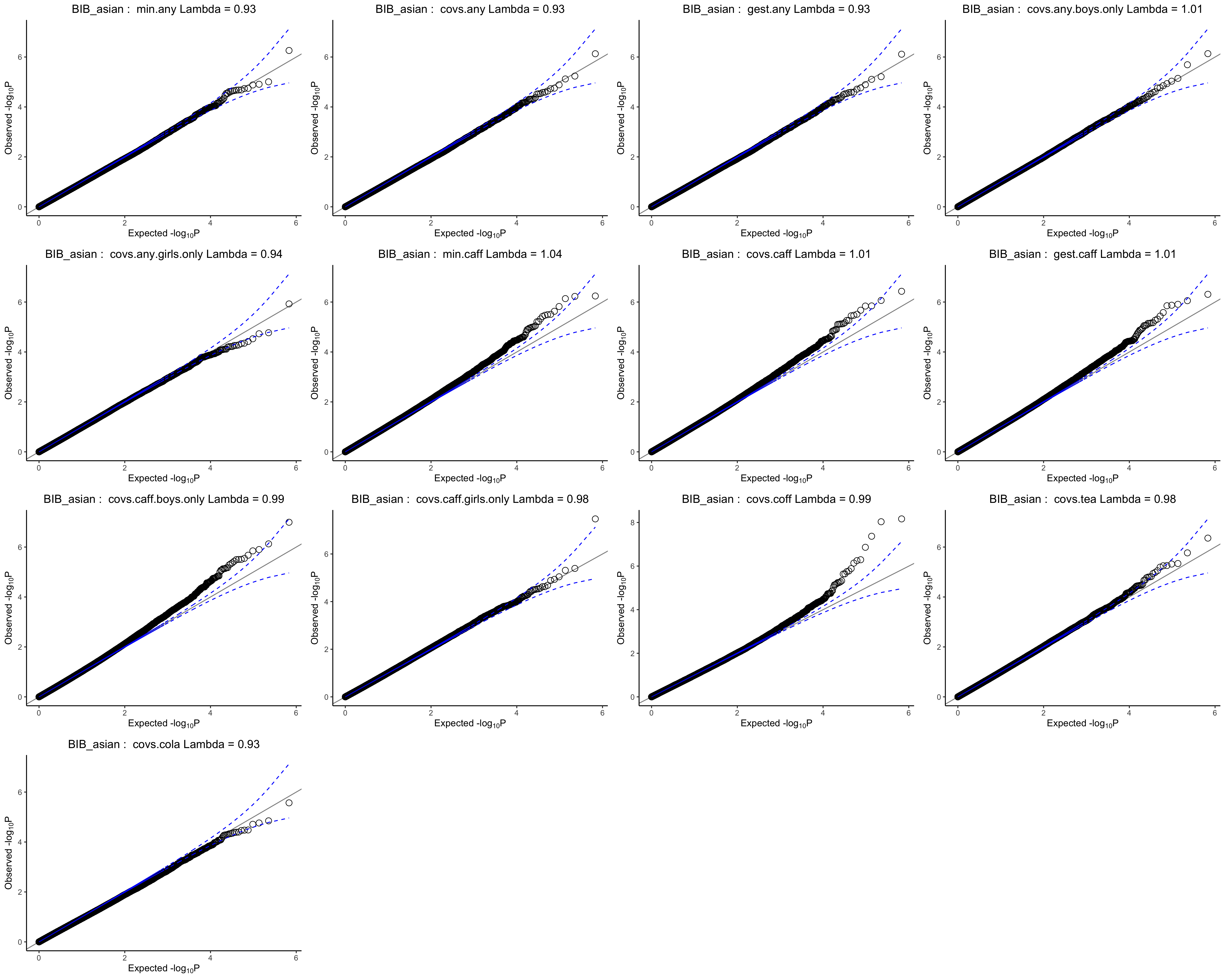

#### Figure S10 BiB (White European) QQ-Plots of caffeine models

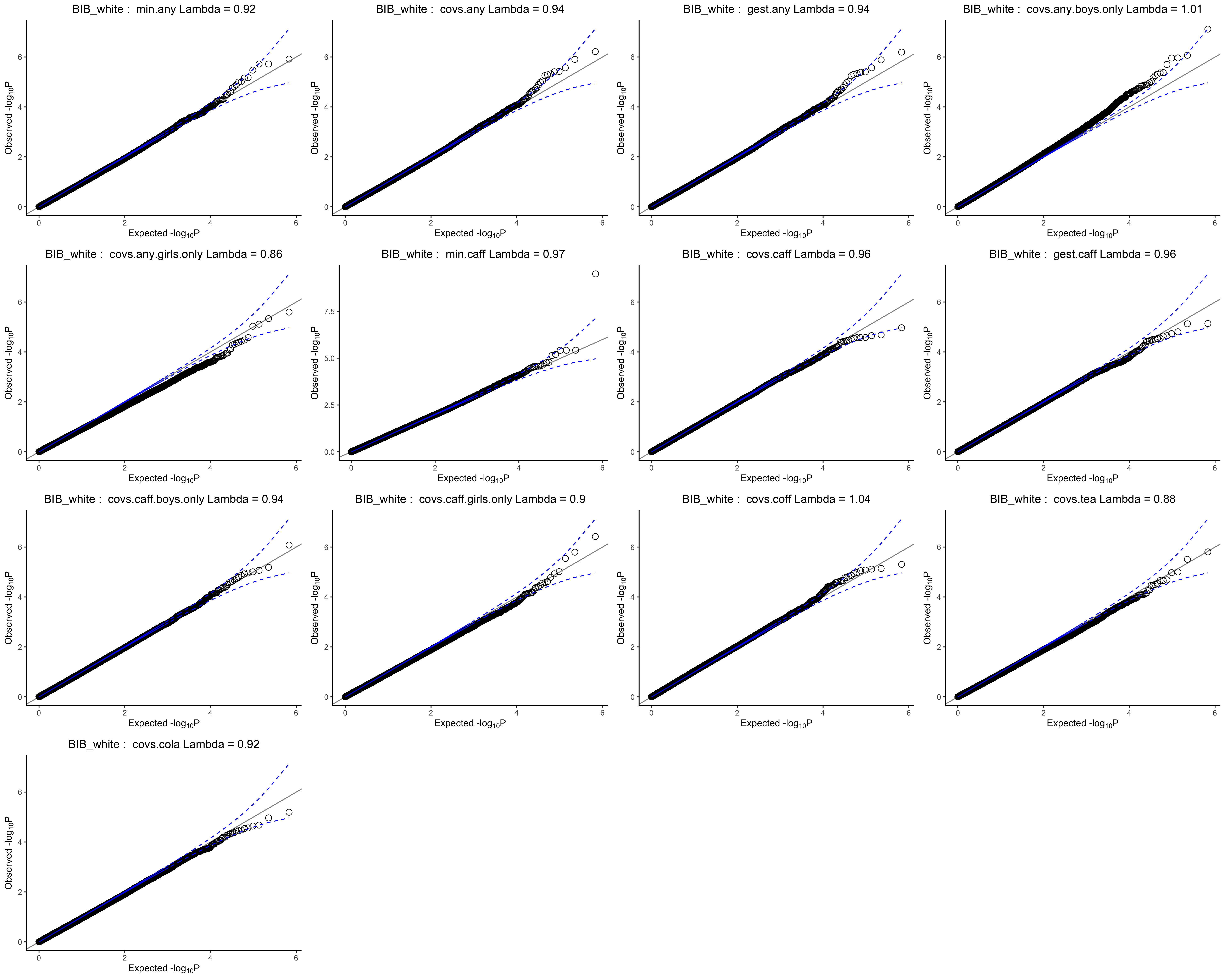

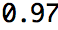

#### Figure S11 Generation R QQ-Plots of caffeine models

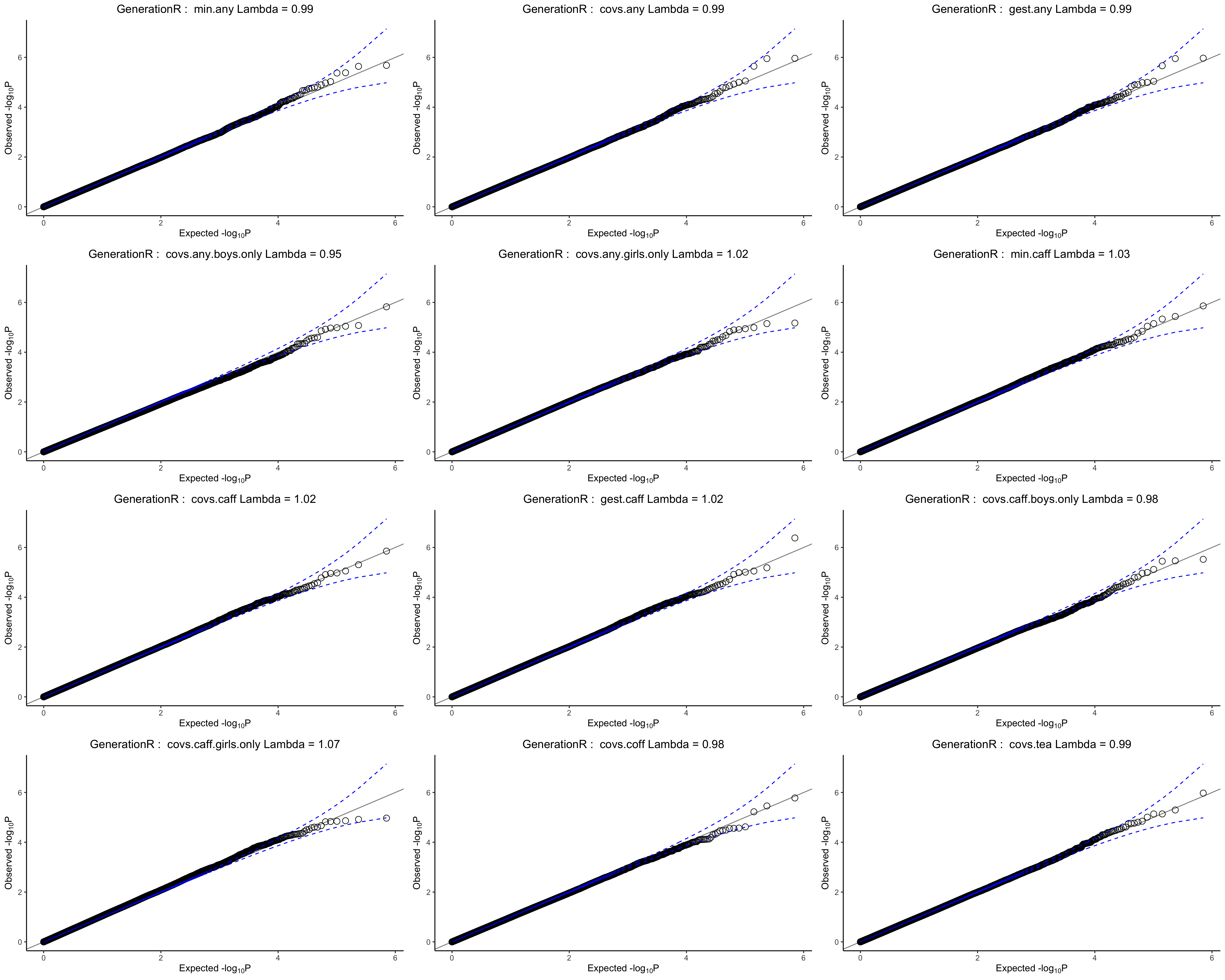

#### Figure S12 MoBa1 QQ-Plots of caffeine models

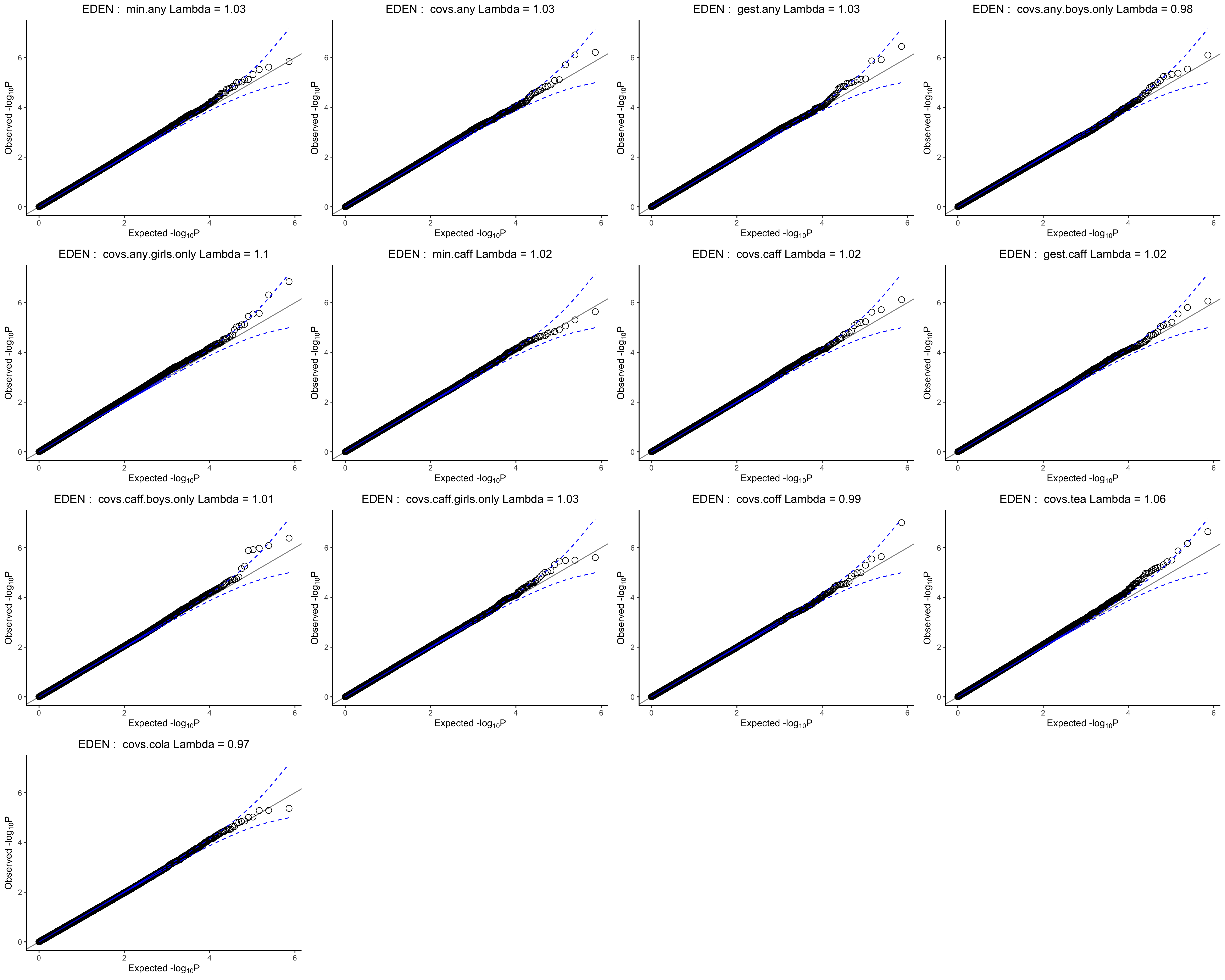

#### Figure S13 EDEN QQ-Plots of caffeine models

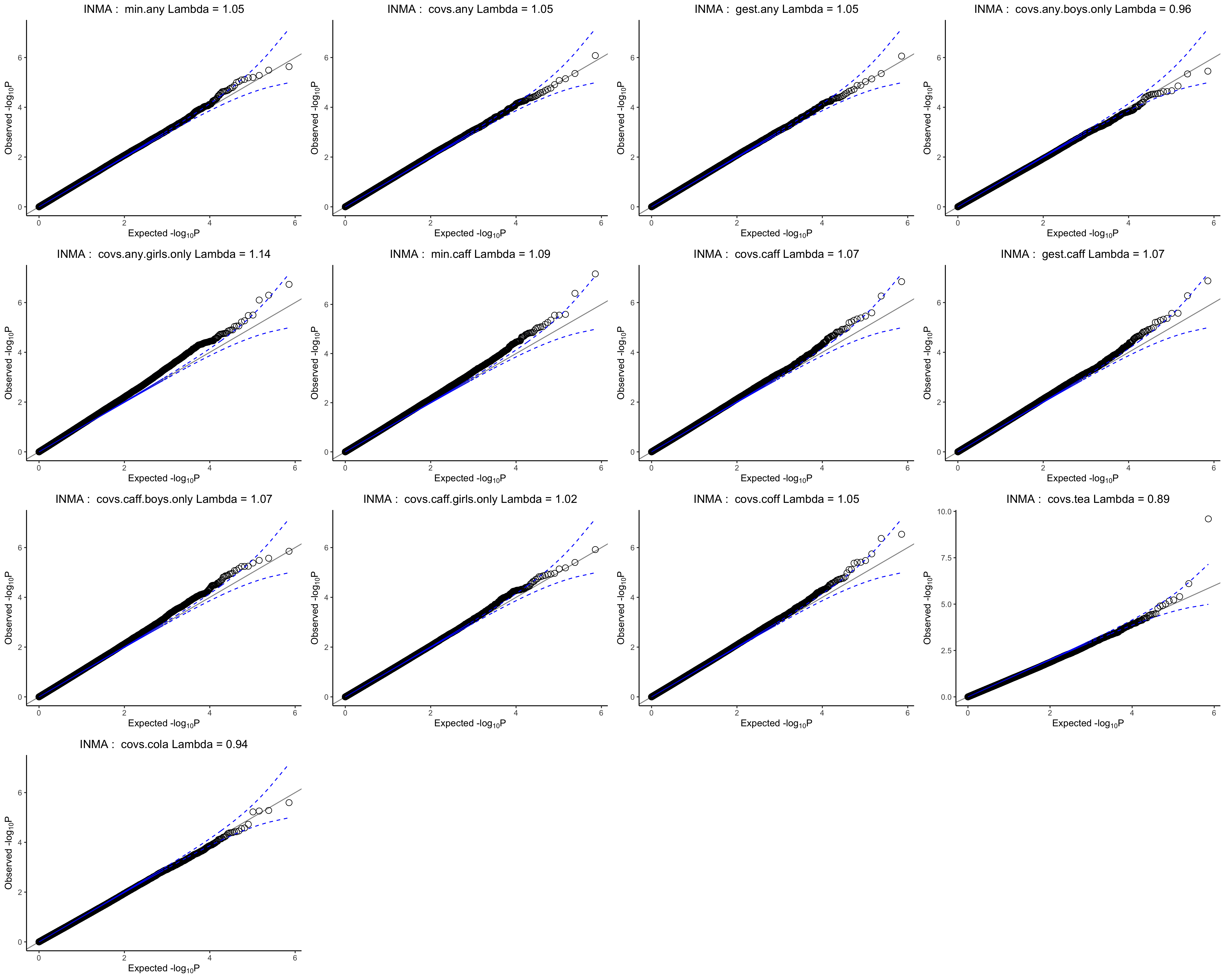

#### Figure S14 INMA QQ-Plots of caffeine models

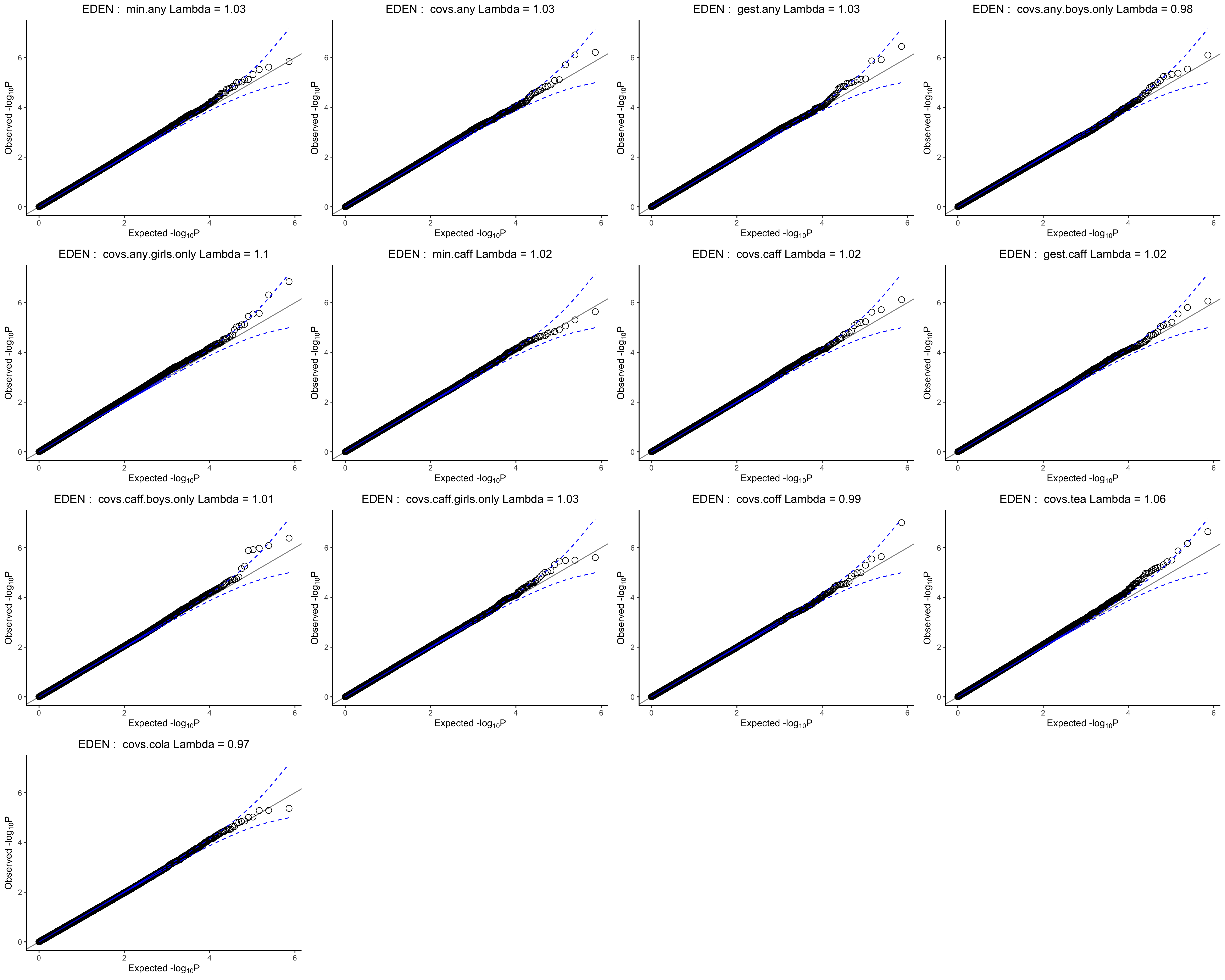

#### Figure S15 Precision plots

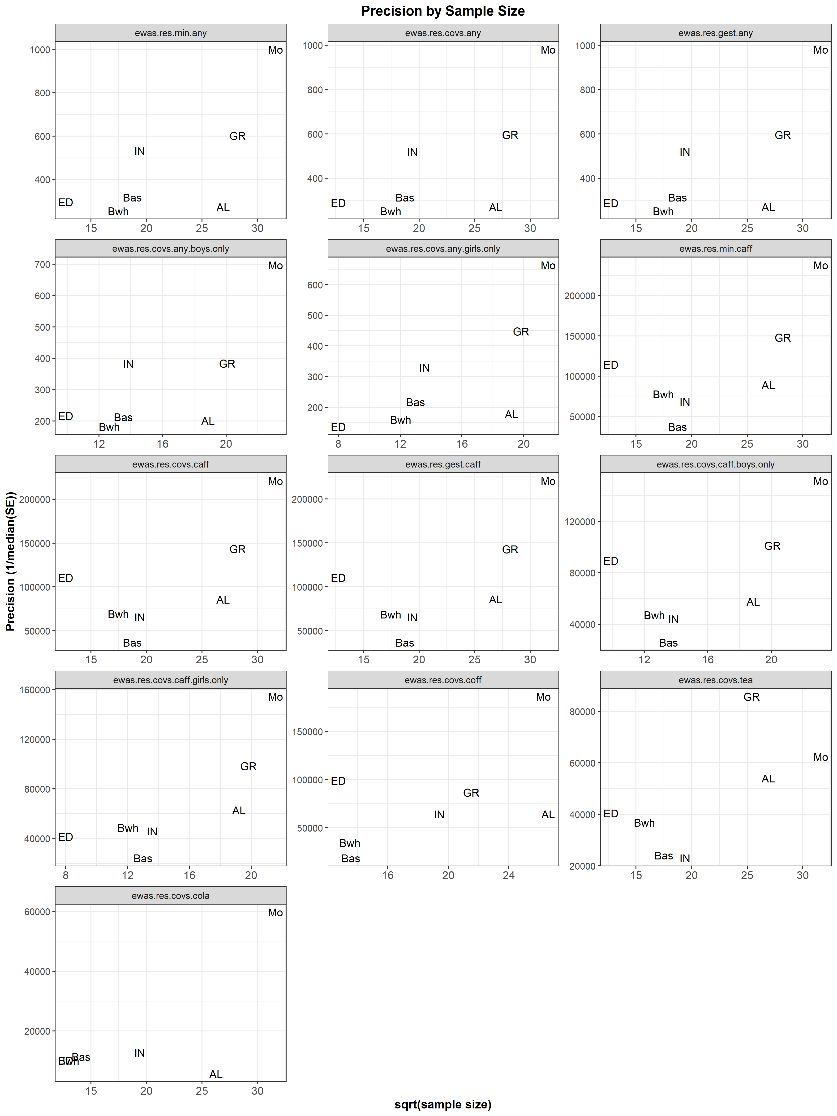

#### Figure S16 Leave-one-out plot of the prenatal caffeine-associated CpG site (Cg19370043)

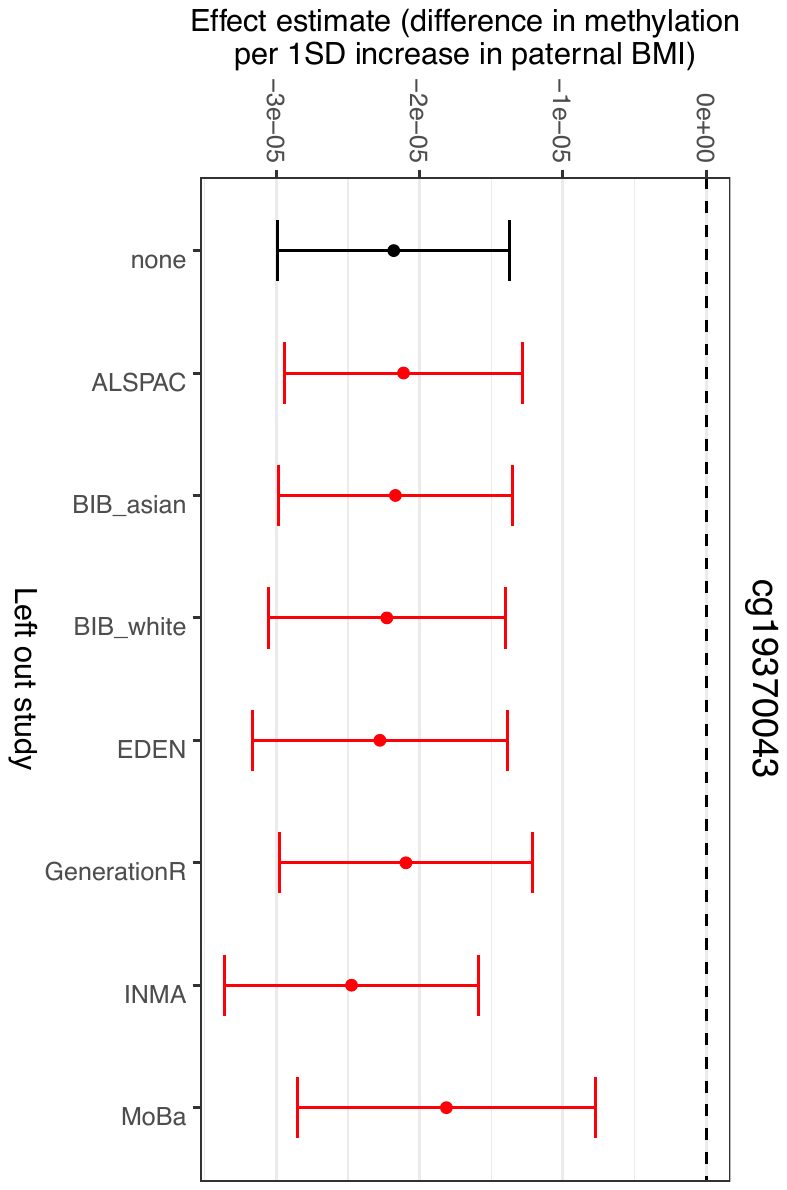

#### Figure S17 Leave-one-out plot of the prenatal cola-associated CpG site (Cg14591243)

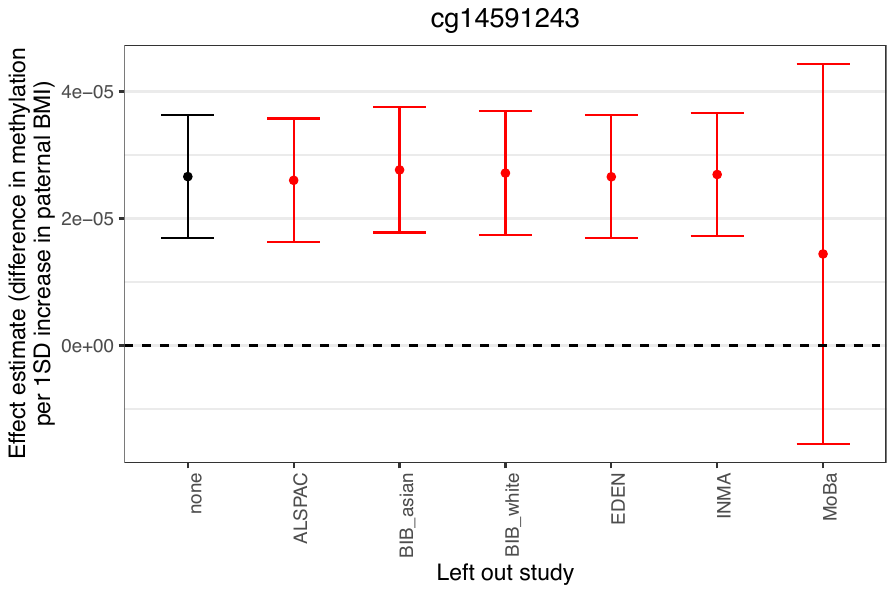

#### Figure S18 Correlation matrix of the meta-analysed caffeine models

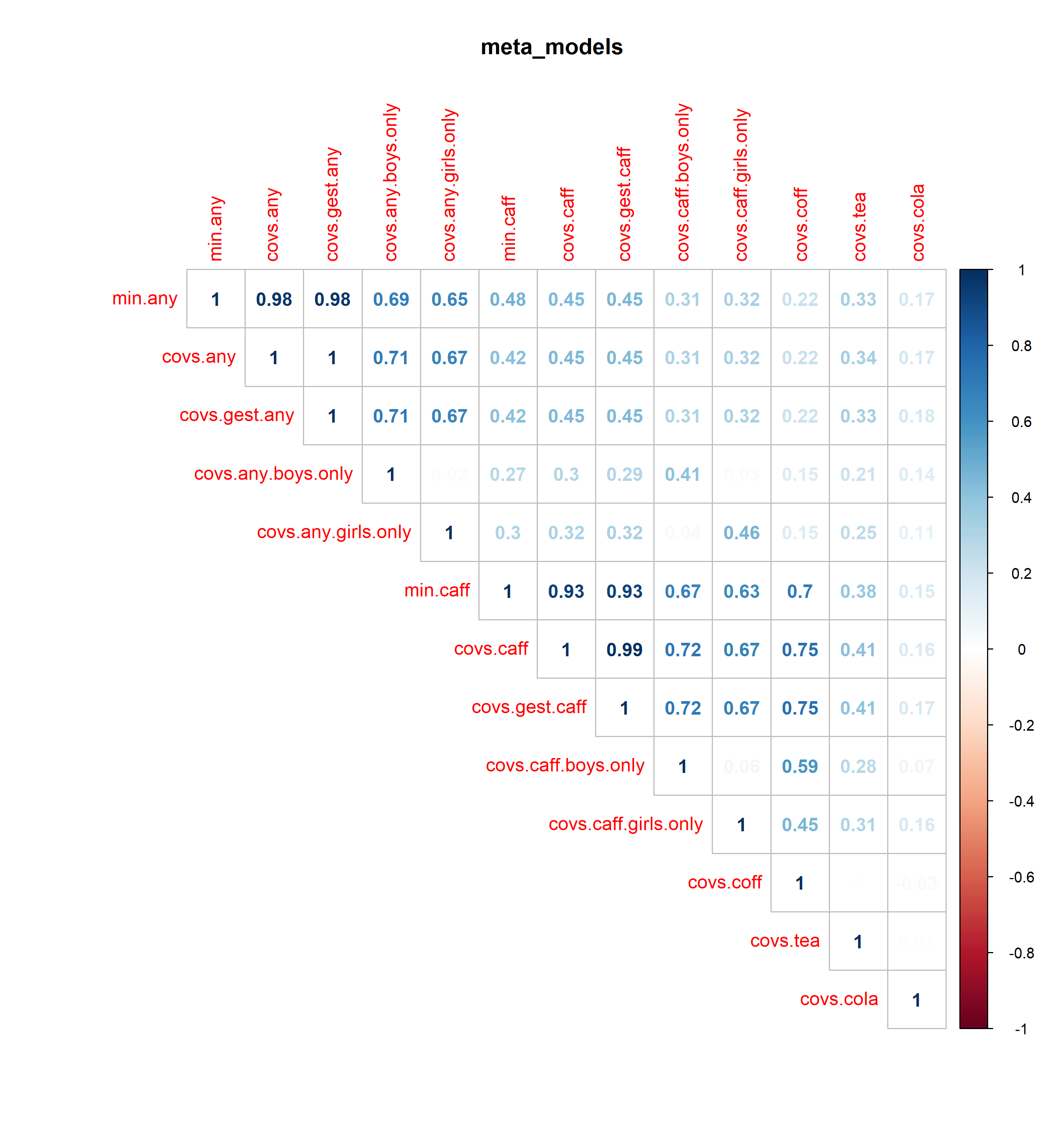

### Differentially methylated regions

#### Table S2 Results from the meta-analysis of differentially methylated regions for total maternal caffeine consumption

| Differentially-methylated region (DMR) | N consecutive CpG sites | Estimate (SE) | Z | P-value | Bonferroni adjusted P-value | Gene |
| --- | --- | --- | --- | --- | --- | --- |
| chr17:58499679-58499911 | 7 | -3.77 x 10^-05^ (5.02 x 10^-06^) | -8.15 | 3.650 x 10^-16^ | 1.420 x 10^-10^ | C17orf64 |
| chr11:6291339-6292490 | 9 | -5.57 x 10^-05^ (7.34 x 10^-06^) | -7.59 | 3.270 x 10^-14^ | 1.270 x 10^-08^ | CCKBR |
| chr12:47219737-47220092 | 10 | -3.77 x 10^-05^ (5.02 x 10^-06^) | -7.52 | 5.680 x 10^-14^ | 2.210 x 10^-08^ | SLC38A4 |
| chr6:30094980-30095341 | 14 | -3.47 x 10^-05^ (4.86 x 10^-06^) | -7.14 | 9.560 x 10^-13^ | 3.720 x 10 ^-07^ | - |
| chr6:33245488-33245770 | 15 | -1.57 x 10^-05^ (2.25 x 10^-06^) | -6.98 | 2.850 x 10^-12^ | 1.110 x 10^-06^ | B3GALT4 |
| chr20:61446962-61447369 | 11 | 2.57 x 10^-05^ (4.12 x 10^-06^) | 6.24 | 4.500 x 10^-10^ | 1.753 x 10^-04^ | COL9A3 |
| chr10:63657059-63657363 | 3 | 6.59 x 10^-05^ (1.08 x 10^-05^) | 6.10 | 1.040 x 10^-09^ | 4.053 x 10 ^-04^ | - |
| chr6:29599160-29599331 | 8 | 4.07 x 10^-05^ (6.70 x 10^-06^) | 6.08 | 1.230 x 10^-09^ | 4.782 x 10^-04^ | GABBR1 |
| chr5:140729653-140730516 | 7 | -2.96 x 10^-05^ (5.01 x 10^-06^) | -5.91 | 3.340 x 10^-09^ | 1.301 x 10^-03^ | PCDHGA2;PCDHGB1;PCDHGA1;PCDHGA3 |
| chr1:117317903-117318185 | 4 | 3.51 x 10^-05^ (5.94 x 10^-06^) | 5.90 | 3.540 x 10^-09^ | 1.380 x 10^-03^ | - |
| chr22:38713874-38714416 | 8 | -2.14 x 10^-05^ (3.77 x 10^-06^) | -5.68 | 1.360 x 10^-08^ | 5.310 x 10 ^-03^ | CSNK1E |
| chr7:130130588-130131258 | 12 | 1.60 x 10^-05^ (2.83 x 10^-06^) | 5.65 | 1.600 x 10^-08^ | 6.223 x 10^-03^ | MESTIT1;MEST |
| chr6:41410759-41411128 | 4 | -1.33 x 10^-05^ (2.40 x 10^-06^) | -5.56 | 2.660 x 10^-08^ | 1.035 x 10^-02^ | - |
| chr7:27153580-27153847 | 6 | -3.66 x 10^-05^ (6.60 x 10^-06^) | -5.55 | 2.810 x 10^-08^ | 1.094 x 10 ^-02^ | HOXA3 |
| chr10:50649723-50650248 | 4 | 4.89 x 10^-05^ (8.84 x 10^-06^) | 5.54 | 3.100 x 10^-08^ | 1.209 x 10 ^-02^ | - |
| chr6:149806131-149806339 | 4 | -3.18 x 10^-05^ (5.74 x 10^-06^) | -5.53 | 3.200 x 10^-08^ | 1.245 x 10 ^-02^ | ZC3H12D |
| chr6:32164723-32165237 | 9 | -2.46 x 10^-05^ (4.47 x 10^-06^) | -5.51 | 3.640 x 10^-08^ | 1.416 x 10^-02^ | GPSM3;NOTCH4 |
| chr4:62383028-62383240 | 3 | -4.34 x 10^-05^ (7.95 x 10^-06^) | -5.46 | 4.790 x 10^-08^ | 1.867 x 10 ^-02^ | LPHN3 |
| chr1:228400217-228400419 | 2 | 4.11 x 10^-05^ (7.66 x 10^-06^) | 5.37 | 8.020^-08^ | 3.125 x 10^-02^ | OBSCN |
| chr6:7468673-7468973 | 4 | -3.68 x 10^-05^ (6.86 x 10^-06^) | -5.36 | 8.340^-08^ | 3.247 x 10 ^-02^ | - |
| chr11:368366-368943 | 15 | -2.03 x 10^-05^ (3.80 x 10^-06^) | -5.34 | 9.250^-08^ | 3.601 x 10 ^-02^ | B4GALNT4 |
| chr7:27142810-27143403 | 8 | 2.96 x 10^-05^ (5.57 x 10^-06^) | 5.32 | 1.060^-07^ | 4.145 x 10 ^-02^ | HOXA2 |

#### Table S3 Results from the meta-analysis of differentially methylated regions for any vs. no maternal caffeine consumption

| Differentially-methylated region (DMR) | N consecutive CpG sites | Estimate (SE) | Z | P-value | Bonferroni adjusted P-value | Gene |
| --- | --- | --- | --- | --- | --- | --- |
| chr6:31734147-31734554 | 10 | 9.44 x 10^-03^ (1.37 x 10^-03^) | 6.91 | 4.975 x 10^-12^ | 1.928 x 10^-06^ | *C6orf27* |
| chr1:11561497-11562134 | 5 | 1.05 x 10^-02^ (1.63 x 10^-03^) | 6.42 | 1.338 x 10^-10^ | 5.184 x 10^-05^ | *PTCHD2* |
| chr22:51016501-51017162 | 8 | 1.16 x 10^-02^ (1.83 x 10^-03^) | 6.32 | 2.691 x 10^-10^ | 1.043 x 10^-04^ | *CPT1B; CHKB-CPT1B* |
| chr17:39969264-39969297 | 3 | -5.99 x 10^-03^ (9.97 x 10^-04^) | -6.01 | 1.894 x 10^-09^ | 7.339 x 10^-04^ | *SC65;FKBP10* |
| chr17:19883326-19883474 | 2 | -1.24 x 10^-02^ (2.13 x 10^-03^) | -5.80 | 6.495 x 10 ^-09^ | 2.517 x 10^-03^ | *-* |
| chr20:61002657-61002866 | 5 | 8.23 x 10^-03^ (1.43^-03^) | 5.75 | 8.997 x 10 ^-09^ | 3.487 x 10 ^-03^ | *C20orf151* |
| chr11:64980819-64981297 | 5 | 8.00 x 10^-03^ (1.44 x 10^-03^) | 5.57 | 2.553 x 10 ^-08^ | 9.893 x 10 ^-03^ | *SLC22A20* |
| chr10:530714-531152 | 6 | 1.08 x 10^-02^ (1.94 x 10^-03^) | 5.55 | 2.885 x 10 ^-08^ | 1.118 x 10 ^-02^ | *DIP2C* |
| chr17:40575479-40575822 | 6 | 3.15 x 10^-03^ (5.70 x 10^-04^) | 5.53 | 3.178 x 10 ^-08^ | 1.232 x 10 ^-02^ | *PTRF* |
| chr9:136567339-136568145 | 3 | 9.21 x 10^-03^ (1.73 x 10^-03^) | 5.34 | 9.508 x 10 ^-08^ | 3.685 x 10 ^-02^ | *SARDH* |
| chr20:61447181-61447490 | 10 | 4.58 x 10^-03^ (8.63 x 10^-04^) | 5.31 | 1.080 x 10 ^-07^ | 4.186 x 10 ^-02^ | *COL9A3* |

#### Table S4 Results from the meta-analysis of differentially methylated regions for maternal coffee consumption

| Differentially-methylated region (DMR) | N consecutive CpG sites | Estimate (SE) | Z | P-value | Bonferroni adjusted P-value | Gene |
| --- | --- | --- | --- | --- | --- | --- |
| chr1:25291385-25292034 | 7 | -4.59 x 10 ^-05^ (5.87 x 10 ^-06^) | -7.83 | 4.831 x 10^-15^ | 1.877 x 10 ^-09^ | *RUNX3* |
| chr22:46449461-46449821 | 5 | -9.14 x 10^-05^ (1.26 x 10 ^-05^) | -7.26 | 3.853 x 10^-13^ | 1.497 x 10 ^-07^ | *C22orf26; LOC150381* |
| chr11:2019732-2020314 | 19 | -2.22 x 10^-05^ (3.33 x 10 ^-06^) | -6.67 | 2.528 x 10^-11^ | 9.824 x 10 ^-06^ | *H19* |
| chr6:49681178-49681742 | 8 | -9.79 x 10^-05^ (1.55 x 10^-05^) | -6.30 | 2.911 x 10^-10^ | 1.131 x 10 ^-04^ | *CRISP2* |
| chr11:2292895-2293173 | 10 | 6.05 x 10^-05^ (1.01 x 10 ^-05^) | 5.98 | 2.212 x 10^-09^ | 8.594 x 10 ^-04^ | *ASCL2* |
| chr1:63249197-63249213 | 4 | 8.92 x 10^-05^(1.60 x 10^-05^) | 5.57 | 2.537 x 10 ^-08^ | 9.859 x 10 ^-03^ | *ATG4C* |
| chr7:5111621-5111916 | 5 | 5.33 x 10 ^-05^ (9.61 x 10 ^-06^) | 5.55 | 2.851 x 10 ^-08^ | 1.108 x 10 ^-02^ | *LOC389458* |
| chr11:368351-368683 | 11 | -4.43 x 10^-05^(8.10 ^-^ x 10^06^) | -5.47 | 4.496 x 10 ^-08^ | 1.747 x 10 ^-02^ | *B4GALNT4* |
| chr17:58499679-58499911 | 7 | -3.93 x 10^-05^(7.20 x 10 ^-06^) | -5.46 | 4.866 x 10^-08^ | 1.891 x 10 ^-02^ | *C17orf64* |
| chr6:155537901-155538055 | 5 | -4.43 x 10 ^-05^(8.20 x 10 ^-06^) | -5.40 | 6.837 x 10 ^-08^ | 2.657 x 10 ^-02^ | *TIAM2* |
| chr2:237478526-237478664 | 3 | -4.98 x 10^-05^(9.24 x 10 ^-06^) | -5.39 | 6.913 x 10 ^-08^ | 2.686 x 10 ^-02^ | *CXCR7* |
| chr2:732577-732961 | 2 | -1.66 x 10^-04^(3.13 x 10^-05^) | -5.30 | 1.169 x 10^-07^ | 4.542 x 10^-02^ | - |

#### Table S5 Results from the meta-analysis of differentially methylated regions for maternal tea consumption

| Differentially-methylated region (DMR) | N consecutive CpG sites | Estimate (SE) | Z | P-value | Bonferroni adjusted P-value | Gene |
| --- | --- | --- | --- | --- | --- | --- |
| chr13:114814024-114814401 | 4 | -1.06 x 10^-04^ (1.54 x 10^-05^) | -6.88 | 6.181 x 10^-12^ | 2.393 x 10^-06^ | *RASA3* |
| chr7:27143046-27143370 | 6 | 6.86 x 10^-05^ (1.04 x 10^-05^) | 6.59 | 4.355 x 10^-11^ | 1.686 x 10^-05^ | *HOXA2* |
| chr6:31127120-31127379 | 7 | 5.47 x 10^-05^ (8.66 x 10^-06^) | 6.31 | 2.766 x 10^-10^ | 1.071 x 10^-04^ | *TCF19; CCHCR1* |
| chr6:160023581-160024002 | 4 | 1.32 x 10^-04^ (2.25 x 10^-05^) | 5.88 | 4.198 x 10^-09^ | 1.625 x 10^-03^ |  |
| chr6:29599160-29599331 | 8 | 6.55 x 10^-05^ (1.12 x 10^-05^) | 5.82 | 5.802 x 10^-09^ | 2.247 x 10^-03^ | *GABBR1* |
| chr11:2019862-2020537 | 18 | 2.10 x 10^-05^ (3.61 x 10^-06^) | 5.81 | 6.068 x 10^-09^ | 2.350 x 10^-03^ | *H19* |
| chr19:51189671-51190179 | 4 | 7.05 x 10^-05^ (1.22 x 10^-05^) | 5.80 | 6.744 x 10^-09^ | 2.611 x 10^-03^ | *SHANK1* |
| chr3:42705828-42706106 | 3 | -6.61 x 10^-05^ (1.15 x 10^-05^) | -5.73 | 1.017 x 10^-08^ | 3.940 x 10^-03^ | *ZBTB47* |
| chr6:136872115-136872119 | 2 | 8.55 x 10^-05^ (1.49 x 10^-05^) | 5.73 | 1.021 x 10^-08^ | 3.955 x 10^-03^ | *MAP7* |
| chr1:117317838-117318185 | 5 | 4.62 x 10^-05^ (8.07 x 10^-06^) | 5.73 | 1.031 x 10^-08^ | 3.992 x 10^-03^ | *-* |
| chr7:155284062-155284759 | 5 | 8.80 x 10^-05^ (1.55 x 10^-05^) | 5.69 | 1.242 x 10^-08^ | 4.811 x 10^-03^ | *-* |
| chr8:144659883-144660772 | 5 | 5.20^-05^ (9.16^-06^) | 5.67 | 1.422 x 10^-08^ | 5.507 x 10^-03^ | *NAPRT1* |
| chr6:32015651-32015737 | 3 | 1.94 x 10^-05^ (3.51 x 10^-06^) | 5.53 | 3.217 x 10^-08^ | 1.246 x 10^-02^ | *TNXB* |
| chr9:136198682-136199172 | 3 | -1.91 x 10^-05^ (3.52 x 10^-06^) | -5.41 | 6.305 x 10^-08^ | 2.442 x 10^-02^ | *SURF6* |
| chr10:63657059-63657363 | 3 | 9.72 x 10^-05^ (1.81 x 10^-05^) | 5.37 | 7.766 x 10^-08^ | 3.007 x 10^-02^ |  |
| chr17:80976690-80977389 | 3 | 2.72 x 10^-05^ (5.12 x 10^-06^) | 5.32 | 1.033 x 10^-07^ | 4.001 x 10^-02^ | *B3GNTL1* |
| chr1:234367145-234367493 | 4 | 1.14 x 10^-04^ (2.15 x 10^-05^) | 5.32 | 1.051 x 10^-07^ | 4.071 x 10^-02^ | *SLC35F3* |
| chr17:58499679-58499816 | 5 | -4.68 x 10^-05^ (8.86 x 10^-06^) | -5.28 | 1.282 x 10^-07^ | 4.966 x 10^-02^ | *C17orf64* |

#### Table S6 Results from the meta-analysis of differentially methylated regions for maternal cola consumption

| Differentially-methylated region (DMR) | N consecutive CpG sites | Estimate (SE) | Z | P-value | Bonferroni adjusted P-value | Gene |
| --- | --- | --- | --- | --- | --- | --- |
| chr3:99979117-99979421 | 3 | -3.48 x 10^-04^ (4.06 x 10^-05^) | -8.57 | 1.024 x 10^-17^ | 4.087 x 10^-12^ | *TBC1D23* |
| chr3:37033632-37033980 | 5 | -1.49 x 10^-03^ (2.06 x 10^-04^) | -7.25 | 4.099 x 10^-13^ | 1.636 x 10^-07^ | *EPM2AIP1;MLH1* |
| chr22:31318103-31318546 | 9 | -4.96 x 10^-03^ (6.99 x 10^-04^) | -7.10 | 1.218 x 10^-12^ | 4.862 x 10^-07^ | *C22orf27* |
| chr2:113992762-113993313 | 7 | -5.24 x 10^-03^ (7.70 x 10^-04^) | -6.80 | 1.032 x 10^-11^ | 4.118 x 10^-06^ | *PAX8* |
| chr1:242220538-242220925 | 3 | -5.53 x 10^-03^ (8.52 x 10^-04^) | -6.49 | 8.403 x 10^-11^ | 3.353 x 10^-05^ |  |
| chr2:11101549-11101592 | 2 | -8.33 x 10^-03^ (1.33 x 10^-03^) | -6.27 | 3.533 x 10^-10^ | 1.410 x 10^-04^ |  |
| chr7:30635762-30636176 | 4 | -4.05 x 10^-03^ (6.56 x 10^-04^) | -6.18 | 6.499 x 10^-10^ | 2.593 x 10^-04^ | *GARS* |
| chr14:50088544-50088598 | 2 | -4.96 x 10^-03^ (8.06 x 10^-04^) | -6.15 | 7.553 x 10^-10^ | 3.014 x 10^-04^ | *RPL36AL;MGAT2* |
| chr7:128530800-128531165 | 2 | 3.21 x 10^-03^ (5.37 x 10^-04^) | 5.97 | 2.332 x 10^-09^ | 9.307 x 10^-04^ | *KCP* |
| chr17:18761852-18761932 | 2 | 2.80 x 10^-04^ (5.05 x 10^-05^) | 5.56 | 2.753 x 10^-08^ | 1.099 x 10^-02^ | *PRPSAP2* |
| chr11:66104174-66104485 | 4 | 1.99 x 10^-03^ (3.59 x 10^-04^) | 5.55 | 2.851 x 10^-08^ | 1.138 x 10^-02^ | *RIN1* |
| chr4:122853963-122854405 | 6 | 5.01 x 10^-03^ (9.20 x 10^-04^) | 5.45 | 5.099 x 10^-08^ | 2.035 x 10^-02^ | *TRPC3* |
| chr13:24519920-24520348 | 3 | -1.23 x 10^-02^ (2.25 x 10^-03^) | -5.45 | 5.124 x 10^-08^ | 2.045 x 10^-02^ |  |
| chr1:152080471-152081002 | 3 | 2.43 x 10^-03^ (4.50 x 10^-04^) | 5.41 | 6.461 x 10^-08^ | 2.578 x 10^-02^ | *TCHH* |

#### Table S7 Results from the meta-analysis of differentially methylated regions for maternal caffeine consumption stratified by female sex

| Differentially-methylated region (DMR) | N consecutive CpG sites | Estimate (SE) | Z | P-value | Bonferroni adjusted P-value | Gene |
| --- | --- | --- | --- | --- | --- | --- |
| chr2:139538222-139539001 | 5 | 2.50 x 10^-03^ (3.44 x 10^-04^) | 7.28 | 3.266 x 10^-13^ | 1.305 x 10^-07^ | *NXPH2* |
| chr19:50931432-50931622 | 4 | 2.08 x 10^-03^ (3.26 x 10^-04^) | 6.38 | 1.812 x 10^-10^ | 7.240 x 10^05^ | *SPIB* |
| chr21:35831996-35832180 | 4 | 1.87 x 10^-03^ (3.05 x 10^-04^) | 6.14 | 8.366 x 10^-10^ | 3.343 x 10^-04^ | *KCNE1* |
| chr4:184930940-184931143 | 4 | 1.32 x 10^-04^ (2.25 x 10^-05^) | 5.85 | 5.048 x 10^-09^ | 2.017 x 10^-03^ | *STOX2* |
| chr6:33257050-33257788 | 18 | 5.78 x 10^-05^  (1.01 x 10^-05^) | 5.74 | 9.618 x 10^-09^ | 3.844 x 10^-03^ | *WDR46* |
| chr11:45715517-45715523 | 2 | -7.31 x 10^-04^ (1.29 x 10^-04^) | -5.65 | 1.627 x 10^-08^ | 6.501 x 10^-03^ |  |
| chr6:168197699-168197921 | 3 | 6.31 x 10^-04^  (1.13 x 10^-04^) | 5.59 | 2.208 x 10^-08^ | 8.823 x 10^-03^ | *C6orf123* |
| chr6:27860893-27860984 | 3 | 2.37 x 10^-04^ (4.34 x 10^-05^) | 5.48 | 4.322 x 10^-08^ | 1.727 x 10^-02^ | *HIST1H2AMHIST1H2BO* |
| chr4:89618637-89618667 | 2 | 1.58 x 10^-03^ (2.90 x 10^-04^) | 5.44 | 5.467 x 10^-08^ | 2.185 x 10^-02^ | *NAP1L5; HERC3* |
| chr20:25565460-25565667 | 2 | -8.45 x 10^-04^ (1.56 x 10^-04^) | -5.41 | 6.327 x 10^-08^ | 2.528 x 10^-02^ | *NINL* |
| chr11:368588-368847 | 6 | -5.26 x 10^-04^ (9.80 x 10^-05^) | -5.37 | 7.859 x 10^-08^ | 3.141 x 10^-02^ | *B4GALNT4* |
| chr4:146018582-146018691 | 2 | -1.08 x 10^-03^ (2.02 x 10^-04^) | -5.37 | 8.036 x 10^-08^ | 3.212 x 10^-02^ | *ANAPC10;ABCE1* |

#### Table S8 Results from the meta-analysis of differentially methylated regions for maternal caffeine consumption stratified by male sex

| Differentially-methylated region (DMR) | N consecutive CpG sites | Estimate (SE) | Z | P-value | Bonferroni adjusted  P-value | Gene |
| --- | --- | --- | --- | --- | --- | --- |
| chr12:58132093-58132558 | 3 | -1.90 x 10^-03^ (2.65 x 10^-04^) | -7.18 | 7.022 x 10^-13^ | 2.837 x 10^-07^ | *AGAP2* |
| chr3:49170599-49171051 | 6 | -6.61 x 10^-04^ (9.84 x 10^-05^) | -6.71 | 1.881 x 10^-11^ | 7.598 x 10^-06^ | *LAMB2* |
| chr7:94286343-94286760 | 16 | 5.06^-^ x 10^04^ (7.56 x 10^-05^) | 6.69 | 2.190 x 10^-11^ | 8.847 x10^-06^ | *SGCE; PEG10* |
| chr13:36871943-36872346 | 9 | 1.01 x 10^-04^ (1.56 x 10^-05^) | 6.44 | 1.219 x 10^-10^ | 4.92 x 10^-05^ | *C13orf38* |
| chr2:3699195-3699563 | 5 | 5.21 x 10^-04^ (8.65 x 10^-05^) | 6.02 | 1.697 x 10^-09^ | 6.856 x 10^-04^ | *-* |
| chr8:95961618-95962383 | 6 | -4.29 x 10^-04^ (7.24 x 10^-05^) | -5.93 | 3.047 x 10^-09^ | 1.231 x 10^-03^ | *TP53INP1* |
| chr19:47852470-47852595 | 3 | 1.36 x 10^-04^ (2.33 x 10^-05^) | 5.86 | 4.637 x 10^-09^ | 1.873 x 10^-03^ | *DHX34* |
| chr6:30614168-30614422 | 7 | 2.51 x 10^-04^ (4.29 x 10^-05^) | 5.85 | 5.019 x 10^-09^ | 2.027 x 10^-03^ | *C6orf136;* |
| chr1:147801103-147801721 | 4 | 2.16 x 10^-03^ (3.79 x 10^-04^) | 5.69 | 1.294 x 10^-08^ | 5.227 x 10^-03^ | *-* |
| chr7:63505768-63506148 | 3 | -1.77 x 10^-03^ (3.12 x 10^-04^) | -5.67 | 1.393 x 10^-08^ | 5.628 x 10^-03^ | *ZNF727* |
| chr7:158110152-158110685 | 4 | 1.21 x 10^-03^ (2.14 x 10^-04^) | 5.66 | 1.551 x 10^-08^ | 6.264 x 10^-03^ | *PTPRN2;* |
| chr11:31391088-31391293 | 3 | 1.89 x 10^-04^ (3.39 x 10^-05^) | 5.58 | 2.378 x 10^-08^ | 9.606 x 10^-03^ | *DCDC1; DNAJC24* |
| chr13:47472050-47472140 | 4 | -5.58 x 10^-04^ (1.04 x 10^-04^) | -5.39 | 6.932 x 10^-08^ | 2.800 x 10^-02^ | *HTR2A* |
| chr7:73894884-73895109 | 3 | -1.79 x 10^-03^ (3.33 x 10^-04^) | -5.39 | 7.224 x 10^-08^ | 2.918 x 10^-02^ | *GTF2IRD1* |
| chr10:23982350-23982387 | 2 | 3.85 x 10^-04^ (7.22 x 10^-05^) | 5.33 | 1.009 x 10^-07^ | 4.075 x 10^-02^ | *KIAA1217* |
| chr11:63974153-63974229 | 3 | 1.55 x 10^-04^ (2.91 x 10^-05^) | 5.32 | 1.018 x 10^-07^ | 4.110 x 10^-02^ | *FERMT3* |
| chr1:20209848-20210269 | 2 | -1.09 x 10^-03^ (2.05 x 10^-04^) | -5.32 | 1.055 x 10^-07^ | 4.261 x 10^-02^ | *OTUD3* |
| chr7:54609587-54609671 | 2 | 2.51 x 10^-04^ (4.72 x 10^-05^) | 5.31 | 1.112 x 10^-07^ | 4.490 x 10^-02^ | *VSTM2A* |

#### Table S9  Crossover of CpG sites and genes of the DMRs of the different caffeine models

|  | **CpGs total caffeine (genes)** | **CpGs any vs. no caffeine (genes)** | **CpGs coffee (genes)** | **CpGs tea**  **(genes)** | **CpGs cola (genes)** | **Total caffeine – Female sex (genes)** | **Total caffeine – Male sex (genes)** |
| --- | --- | --- | --- | --- | --- | --- | --- |
| **CpGs total caffeine (genes)** | 167 CpGs (100%) | 7 CpGs (*COL9A3*), crossover = 3% | 17 CpGs (*C17orf64; B4GALNT4*),  crossover = 7% | 26 CpGs (*C17orf64 GABBR1; HOXA2*), crossover = 11% | 0 CpGs  (0 genes) | 6 CpGs (*B4GALNT4*), crossover = 3% | 0 CpGs  (0 genes) |
| - | **CpGs any vs. no caffeine (genes)** | 63 CpGs  (100%) | 0 CpGs  (0 genes) | 0 CpGs  (0 genes) | 0 CpGs  (0 genes) | 0 CpGs (0 genes) | 0 CpGs  (0 genes) |
| - | - | **CpGs coffee (genes)** | 86 CpGs  (100%) | 19 CpGs  (*H19; C17orf64*), crossover = 12% | 0 CpGs  (0 genes) | 4 CpGs (*B4GALNT4*), crossover = 3% | 0 CpGs  (0 genes) |
| - | - | - | **CpGs tea (genes)** | 92 CpGs  (100%) | 0 CpGs  (0 genes) | 0 CpGs  (0 genes) | 0 CpGs  (0 genes) |
| - | - | - | - | **CpGs cola**  **(genes)** | 55 CpGs  (100%) | 0 CpGs  (0 genes) | 0 CpGs  (0 genes) |
| - | - | - | - | - | **Total caffeine – Female sex (genes)** | 55 CpGs  (100%) | 0 CpGs  (0 genes) |
| - | - | - | - | - | - | **Total caffeine – Male sex (genes)** | 85 CpGs (100%) |
| *Note. Cells highlighted in blue represent the total number of CpG sites (CpGs) that were contained across the DMRs of each caffeine model. Darker shading of blue = more overlap. Crossover = percentage of crossover CpG sites between the different models (N CpGs crossover / (N CpGs model 1 + N CpGs model 2 – N CpGs crossover).* | | | | | | | |

#### Table S10 Top 5 GO terms and KEGG pathways for CpGs in DMRs (BP = biological process; MF = molecular function; CC = cell compartment)

| **Model** | **Ontology/KEGG** | **Term/Pathway** | **ID** | **N CpGs differentially methylated** | **N CpGs in term/pathway** | **P-value for enrichment** | **FDR corrected P-value** |
| --- | --- | --- | --- | --- | --- | --- | --- |
| **Total caffeine** |  |  |  |  |  |  |  |
|  | BP | cell-cell adhesion via plasma-membrane adhesion molecules | GO:0098742 | 5 | 260 | 0.0001 | 1 |
|  | BP | homophilic cell adhesion via plasma membrane adhesion molecules | GO:0007156 | 4 | 160 | 0.0002 | 1 |
|  | MF | calcium ion binding | GO:0005509 | 6 | 670 | 0.0006 | 1 |
|  | CC | integral component of plasma membrane | GO:0005887 | 8 | 1520 | 0.001 | 1 |
|  | CC | intrinsic component of plasma membrane | GO:0031226 | 8 | 1595 | 0.002 | 1 |
|  | KEGG | Glycosphingolipid biosynthesis - ganglio series | path:hsa00604 | 1 | 15 | 0.033 | 1 |
|  | KEGG | Neuroactive ligand-receptor interaction | path:hsa04080 | 2 | 317 | 0.048 | 1 |
|  | KEGG | Various types of N-glycan biosynthesis | path:hsa00513 | 1 | 37 | 0.052 | 1 |
|  | KEGG | Circadian rhythm | path:hsa04710 | 1 | 30 | 0.055 | 1 |
|  | KEGG | Hippo signaling pathway - multiple species | path:hsa04392 | 1 | 28 | 0.073 | 1 |
| **Any vs. no Caffeine** |  |  |  |  |  |  |  |
|  | MF | sarcosine dehydrogenase activity | GO:0008480 | 1 | 1 | 0.001 | 1 |
|  | BP | sarcosine catabolic process | GO:1901053 | 1 | 1 | 0.001 | 1 |
|  | MF | rRNA primary transcript binding | GO:0042134 | 1 | 1 | 0.001 | 1 |
|  | MF | oxidoreductase activity, acting on the CH-NH group of donors, flavin as acceptor | GO:0046997 | 1 | 2 | 0.001 | 1 |
|  | CC | endoplasmic reticulum | GO:0005783 | 5 | 1287 | 0.0010 | 1 |
|  | KEGG | Glycine, serine and threonine metabolism | path:hsa00260 | 1 | 35 | 0.019 | 1 |
|  | KEGG | Fatty acid degradation | path:hsa00071 | 1 | 41 | 0.024 | 1 |
|  | KEGG | Fatty acid metabolism | path:hsa01212 | 1 | 55 | 0.041 | 1 |
|  | KEGG | PPAR signaling pathway | path:hsa03320 | 1 | 72 | 0.045 | 1 |
|  | KEGG | Adipocytokine signaling pathway | path:hsa04920 | 1 | 66 | 0.051 | 1 |
| **Coffee** |  |  |  |  |  |  |  |
|  | BP | mesenchymal stem cell migration | GO:1905319 | 1 | 3 | 0.002 | 1 |
|  | BP | regulation of mesenchymal stem cell migration | GO:1905320 | 1 | 3 | 0.002 | 1 |
|  | BP | positive regulation of mesenchymal stem cell migration | GO:1905322 | 1 | 3 | 0.002 | 1 |
|  | MF | C-X-C chemokine binding | GO:0019958 | 1 | 6 | 0.003 | 1 |
|  | MF | N-acetyl-beta-glucosaminyl-glycoprotein 4-beta-N-acetylgalactosaminyltransferase activity | GO:0033842 | 1 | 2 | 0.003 | 1 |
|  | KEGG | Autophagy - other | path:hsa04136 | 1 | 29 | 0.019 | 1 |
|  | KEGG | Various types of N-glycan biosynthesis | path:hsa00513 | 1 | 37 | 0.023 | 1 |
|  | KEGG | Viral protein interaction with cytokine and cytokine receptor | path:hsa04061 | 1 | 95 | 0.031 | 1 |
|  | KEGG | Th1 and Th2 cell differentiation | path:hsa04658 | 1 | 87 | 0.063 | 1 |
|  | KEGG | Autophagy - animal | path:hsa04140 | 1 | 129 | 0.081 | 1 |
| **Tea** |  |  |  |  |  |  |  |
|  | MF | nicotinate phosphoribosyltransferase activity | GO:0004516 | 1 | 1 | 0.0004 | 1 |
|  | BP | nicotinate nucleotide biosynthetic process | GO:0019357 | 1 | 1 | 0.0004 | 1 |
|  | BP | nicotinate nucleotide salvage | GO:0019358 | 1 | 1 | 0.0004 | 1 |
|  | BP | pyridine nucleotide salvage | GO:0019365 | 1 | 1 | 0.0004 | 1 |
|  | BP | NAD salvage | GO:0034355 | 1 | 1 | 0.0004 | 1 |
|  | KEGG | Nicotinate and nicotinamide metabolism | path:hsa00760 | 1 | 34 | 0.022 | 1 |
|  | KEGG | Taste transduction | path:hsa04742 | 1 | 82 | 0.066 | 1 |
|  | KEGG | GnRH secretion | path:hsa04929 | 1 | 62 | 0.098 | 1 |
|  | KEGG | ECM-receptor interaction | path:hsa04512 | 1 | 86 | 0.119 | 1 |
|  | KEGG | GABAergic synapse | path:hsa04727 | 1 | 84 | 0.125 | 1 |
| **Cola** |  |  |  |  |  |  |  |
|  | MF | alpha-1,6-mannosylglycoprotein 2-beta-N-acetylglucosaminyltransferase activity | GO:0008455 | 1 | 1 | 0.0003 | 1 |
|  | BP | thyroid-stimulating hormone secretion | GO:0070460 | 1 | 1 | 0.002 | 1 |
|  | BP | regulation of thyroid-stimulating hormone secretion | GO:2000612 | 1 | 1 | 0.002 | 1 |
|  | CC | late recombination nodule | GO:0005715 | 1 | 1 | 0.002 | 1 |
|  | BP | meiotic metaphase I plate congression | GO:0043060 | 1 | 1 | 0.002 | 1 |
|  | MF | alpha-1,6-mannosylglycoprotein 2-beta-N-acetylglucosaminyltransferase activity | GO:0008455 | 1 | 1 | 0.0003 | 1 |
|  | KEGG | Mismatch repair | path:hsa03430 | 1 | 22 | 0.018 | 1 |
|  | KEGG | Aminoacyl-tRNA biosynthesis | path:hsa00970 | 1 | 43 | 0.029 | 1 |
|  | KEGG | Various types of N-glycan biosynthesis | path:hsa00513 | 1 | 37 | 0.032 | 1 |
|  | KEGG | N-Glycan biosynthesis | path:hsa00510 | 1 | 48 | 0.036 | 1 |
|  | KEGG | Fanconi anemia pathway | path:hsa03460 | 1 | 48 | 0.037 | 1 |
| **Total caffeine – female sex** |  |  |  |  |  |  |  |
|  | CC | nucleosome | GO:0000786 | 2 | 90 | 0.0017 | 1 |
|  | MF | endoribonuclease inhibitor activity | GO:0060698 | 1 | 2 | 0.002 | 1 |
|  | BP | negative regulation of ribonuclease activity | GO:0060701 | 1 | 2 | 0.002 | 1 |
|  | BP | negative regulation of endoribonuclease activity | GO:0060702 | 1 | 2 | 0.002 | 1 |
|  | CC | DNA packaging complex | GO:0044815 | 2 | 98 | 0.002 | 1 |
|  | KEGG | Systemic lupus erythematosus | path:hsa05322 | 2 | 113 | 0.003 | 0.93 |
|  | KEGG | Ubiquitin mediated proteolysis | path:hsa04120 | 2 | 132 | 0.006 | 0.93 |
|  | KEGG | Alcoholism | path:hsa05034 | 2 | 164 | 0.008 | 0.93 |
|  | KEGG | Various types of N-glycan biosynthesis | path:hsa00513 | 1 | 37 | 0.033 | 1 |
|  | KEGG | Progesterone-mediated oocyte maturation | path:hsa04914 | 1 | 86 | 0.078 | 1 |
| **Total caffeine – male sex** |  |  |  |  |  |  |  |
|  | BP | positive regulation of phosphatidylinositol biosynthetic process | GO:0010513 | 2 | 5 | 1.2 x 10^-05^ | 0.275 |
|  | BP | regulation of phosphatidylinositol biosynthetic process | GO:0010511 | 2 | 7 | 4.2 x10 ^-05^ | 0.480 |
|  | BP | positive regulation of phospholipid biosynthetic process | GO:0071073 | 2 | 11 | 7.2 x 10^-05^ | 0.544 |
|  | BP | regulation of phospholipid biosynthetic process | GO:0071071 | 2 | 18 | 2.4 10^-04^ | 1.000 |
|  | BP | response to methyl methanesulfonate | GO:0072702 | 1 | 1 | 6.9 x 10^-04^ | 1.000 |

### Cohort-specific acknowledgments

##### ALSPAC

We are extremely grateful to all the families who took part in this study, the midwives for their help in recruiting them, and the whole ALSPAC team, which includes interviewers, computer and laboratory technicians, clerical workers, research scientists, volunteers, managers, receptionists and nurses.

##### BIB

Born in Bradford is only possible because of the enthusiasm and commitment of the Children and Parents in BiB. We are grateful to all the participants, practitioners and researchers who have made Born in Bradford happen. 450K DNAm and genotype array data was generated in the Bristol Bioresource Laboratory Illumina Facility, University of Bristol.

##### Generation R

The Generation R Study is conducted by Erasmus MC, University Medical Center Rotterdam, in close collaboration with the School of Law and Faculty of Social Sciences of the Erasmus University Rotterdam, the Municipal Health Service Rotterdam area, Rotterdam, the Rotterdam Homecare Foundation, Rotterdam and the Stichting Trombosedienst & Artsenlaboratorium Rijnmond (STAR-MDC), Rotterdam. We gratefully acknowledge the contribution of children and parents, general practitioners, hospitals, midwives and pharmacies in Rotterdam. The study protocol was approved by the Medical Ethical Committee of the Erasmus Medical Centre, Rotterdam. Written informed consent was obtained for all participants. The generation and management of the Illumina 450K methylation array data (EWAS data) for the Generation R Study was executed by the Human Genotyping Facility of the Genetic Laboratory of the Department of Internal Medicine, Erasmus MC, the Netherlands. We thank Mr. Michael Verbiest, Ms. Mila Jhamai, Ms. Sarah Higgins, Mr. Marijn Verkerk and Dr. Lisette Stolk for their help in creating the EWAS database. We thank Dr. A.Teumer for his work on the quality control and normalization scripts.

##### MoBa

We are grateful to all the participating families in Norway who take part in this on-going cohort study.

##### INMA

INMA researchers would like to thank all the participants for their generous collaboration. A full roster of the INMA Project Investigators can be found at <https://www.proyectoinma.org/proyecto-inma/investigadores/>

##### EDEN

We are indebted to the EDEN participating families and the midwife research assistants (L. Douhaud, S. Bedel, B. Lortholary, S. Gabriel, M. Rogeon, M. Malinbaum) for data collection. We thank the EDEN mother-child cohort study group (I. Annesi-Maesano, J.Y Bernard, J. Botton, M.A. Charles, P. Dargent-Molina, B. de Lauzon-Guillain, P. Ducimetière, M. de Agostini, B. Foliguet, A. Forhan, X. Fritel, A. Germa, V. Goua, R. Hankard, B. Heude, M. Kaminski, B. Larroque†, N. Lelong, J. Lepeule, G. Magnin, L. Marchand, C. Nabet, F. Pierre, R. Slama, M.J. Saurel-Cubizolles, M. Schweitzer, O. Thiebaugeorges).

### Cohort-specific funding statements

##### ALSPAC

The UK Medical Research Council and Wellcome (Grant ref: 217065/Z/19/Z) and the University of Bristol provide core support for ALSPAC. This publication is the work of the authors and Laura Schellhas will serve as guarantors for the contents of this paper. A comprehensive list of grants funding is available on the ALSPAC website (http://www.bristol.ac.uk/alspac/external/documents/grant-acknowledgements.pdf).

This research was performed in the UK Medical Research Council Integrative Epidemiology Unit (grant number: MC_UU_00011/7 and MC_UU_00011/5) and also supported by the National Institute for Health Research (NIHR) Bristol Biomedical Research Centre at University Hospitals Bristol NHS Foundation Trust and the University of Bristol.

This research was also conducted as part of the CAPICE (Childhood and Adolescence Psychopathology: unravelling the complex etiology by a large Interdisciplinary Collaboration in Europe) project, funded by the European Union’s Horizon 2020 research and innovation programme, Marie Sklodowska Curie Actions – MSCA-ITN-2016 – Innovative Training Networks under grant agreement number 721567. This study was supported by the NIHR Biomedical Research Centre at the University Hospitals Bristol NHS Foundation Trust and the University of Bristol. The views expressed in this publication are those of the authors and not necessarily those of the NHS, the

National Institute for Health Research or the Department of Health and Social Care.

GCS is financially supported by an MRC New Investigator Research (grant code MR/S009310/1), an MRC project grant (MR/W020297/1) and the European Joint Programming Initiative “A Healthy Diet for a Healthy Life” (JPI HDHL, NutriPROGRAM project, UK MRC MR/S036520/1)

##### Born in Bradford (BIB)

BiB receives core funding from the Wellcome Trust (WT101597MA), the British Heart Foundation (CS/16/4/32482), a joint grant from the UK Medical Research Council (MRC) and UK Economic and Social Science Research Council (ESRC) (MR/N024397/1) and the National Institute for Health Research (NIHR) under its Collaboration for Applied Health Research and Care (CLAHRC) for Yorkshire and Humber. The research presented in this paper, including obtaining genome-wide and epigenome-wide DNAm data is supported by the US National Institute of Health (R01 DK10324) and European Research Council under the European Union’s Seventh Framework Programme (FP7/2007-2013) / ERC grant agreement no 669545.

##### Generation R

The general design of the Generation R Study is made possible by financial support from the Erasmus MC, Erasmus University Rotterdam, the Netherlands Organization for Health Research and Development and the Ministry of Health, Welfare and Sport. The EWAS data were funded by a grant to VWJ from the Netherlands Genomics Initiative (NGI)/Netherlands Organisation for Scientific Research (NWO) Netherlands Consortium for Healthy Aging (NCHA; project nr. 050-060-810), by funds from the Genetic Laboratory of the Department of Internal Medicine, Erasmus MC, and by a grant from the National Institute of Child and Human Development (R01HD068437). V.W.J. received a Consolidator Grant from the European Research Council (ERC-2014-CoG-648916). This project received funding from the European Union’s Horizon 2020 research and innovation programme (733206, LifeCycle; 874739, LongITools; 874583, ATHLETE; 824989, EUCAN-Connect) and from the European Joint Programming Initiative “A Healthy Diet for a Healthy Life” (JPI HDHL, NutriPROGRAM project, ZonMw the Netherlands no.529051022 and PREcisE project ZonMw the Netherlands no.529051023).

##### MoBa

The Norwegian Mother, Father and Child Cohort Study is supported by the Norwegian Ministry of Health and Care Services and the Ministry of Education and Research. For this work, MoBa 1 and 2 were supported by the Intramural Research Program of the NIH, National Institute of Environmental Health Sciences (Z01-ES-49019) and the Norwegian Research Council/BIOBANK (grant no 221097). This work was partly supported by the Research Council of Norway through its Centres of Excellence funding scheme, project number 262700. MoBa3 epigenomics data analyses were funded by INCA/Plan Cancer-EVA-INSERM, France, and the International Childhood Cancer Cohort Consortium (I4C), and performed by the Epigenetics Group at the International Agency for Research on Cancer (IARC, Lyon, France). Where authors are identified as personnel of the International Agency for Research on Cancer / World Health Organization, the authors alone are responsible for the views expressed in this article and they do not necessarily represent the decisions, policy or views of the International Agency for Research on Cancer / World Health Organization.

##### INMA

This study was funded by grants from Instituto de Salud Carlos III (Red INMA G03/176; CB06/02/0041; PI041436; PI081151 incl. FEDER funds), Generalitat de Catalunya-CIRIT 1999SGR 00241, Fundació La marató de TV3 (090430), EU Commission (261357-MeDALL: Mechanisms of the Development of ALLergy), European Research Council (268479-BREATHE: BRain dEvelopment and Air polluTion ultrafine particles in scHool children, and the European Joint Programming Initiative “A Healthy Diet for a Healthy Life” (JPI HDHL and  Instituto de Salud Carlos III) under the grant agreement no AC18/00006 (NutriPROGRAM project). We acknowledge support from the Spanish Ministry of Science and Innovation and the State Research Agency through the “Centro de Excelencia Severo Ochoa 2019-2023” Program (CEX2018-000806-S), and support from the Generalitat de Catalunya through the CERCA Program.

##### EDEN

Foundation for medical research (FRM), National Agency for Research (ANR), National Institute for Research in Public health (IRESP: TGIR cohorte santé 2008 program), French Ministry of Health (DGS), French Ministry of Research, INSERM Bone and Joint Diseases National Research (PRO-A) and Human Nutrition National Research Programs, Paris–Sud University, Nestlé, French National Institute for Population Health Surveillance (InVS), French National Institute for Health Education (INPES), the European Union FP7 programs (FP7/2007-2013, HELIX, ESCAPE, ENRIECO, Medall projects), Diabetes National Research Program (in collaboration with the French Association of Diabetic Patients (AFD), French Agency for Environmental Health Safety (now ANSES), Mutuelle Générale de l’Education Nationale complementary health insurance (MGEN), French national agency for food security, French speaking association for the study of diabetes and metabolism (ALFEDIAM), grant # 2012/51290-6 Sao Paulo Research Foundation (FAPESP), EU funded MeDALL project.
